# Transcranial Magnetic Stimulation Depression Treatment Induces Positive Bias in Task-Related Brain Function: Results From the BRAEN-MAP Trial

**DOI:** 10.1101/2025.01.13.25320448

**Authors:** Verena Sarrazin, Paulo Suen, Beatriz Cavendish, Marieke Martens, Pedro Henrique Rodrigues da Silva, Anne Britto, Matheus Rassi, Mariana Baptista, Andre R Brunoni, Jacinta O’Shea

## Abstract

Antidepressant treatments are theorised to act by inducing a positive bias in information processing early on during treatment. Here, we tested for the first time whether this theory generalises to transcranial magnetic stimulation (TMS) treatment, an effective therapy for treatment-resistant depression. 49 patients with major depression received 20 sessions of open-label intermittent theta-burst stimulation applied to left dorsolateral prefrontal cortex. At baseline and after eight stimulation sessions, positive bias was assessed using behavioural and functional magnetic resonance imaging tasks presenting emotional faces. Clinical improvement at the end of treatment was related to an early increase in positive bias (1) in misclassification of emotional faces, (2) in the response of the default mode network (DMN) to emotional faces including rostral anterior cingulate cortex (ACC), and (3) in connectivity between rostral ACC and DMN. These neural changes predicted clinical improvement at the end of treatment beyond early symptom reduction. The results suggest that TMS treatment increases positive bias early on during treatment, and that the neural mechanisms might differ from that of antidepressant drugs.

## 1 Introduction

Depression has been identified by the World Health Organisation as the largest contributor to global disability [1]. Around 30% of patients do not respond to conventional treatments such as medication [2, 3]. Transcranial magnetic stimulation (TMS) applied to left dorsolateral prefrontal cortex (DLPFC) is a promising new treatment which has been shown to be effective for more than half of the patients who did not respond to conventional treatments [4–6]. However, little is known about the cognitive mechanism of action of TMS treatment.

Antidepressant treatments are theorised to act by increasing the focus on positive rather than negative information processing early on during treatment [7–9]. Increased processing of positive information is theorised to counteract negative biases (i.e. excessive focus on negative information) which cause and maintain symptoms of depression [10]. While the neuropsychological theory of antidepressant action was developed based on evidence from drug treatments, increased positive bias has been proposed as a common mechanism of action shared across different treatment modalities [10]. To date, this theory has not directly been tested for TMS treatment.

Negative biases in depression are hypothesised to arise from increased activity in subcortical regions (such as amygdala and subgenual cingulate) in response to negative stimuli, accompanied by decreased top-down control from DLPFC [10]. Neuroimaging studies suggest that TMS depression treatment might normalise interaction within this cortico-limbic circuit, however, these studies were conducted at rest in the absence of cognitive tasks [11–13]. Preliminary evidence from task-based studies suggests that a single session of TMS might increase positive bias, for example, as reflected in increased activity in frontal regions in response to positive stimuli [14], decreased subcortical response to negative stimuli [14, 15], and improved memory retrieval for positive stimuli [16, 17].

The key argument of the neuropsychological theory is that early changes in information processing *precede* and *cause* clinical improvement. However, previous studies on TMS treatment compared measures acquired at baseline to those acquired at the end of treatment (i.e. once patients have recovered)(e.g. [12]). To investigate potential mechanisms of action that drive clinical improvement over time, it is necessary to assess changes *early on* during the treatment (see [18]).

To the best of our knowledge, this study investigated for the first time how positive bias in the processing of emotional stimuli changes early on during TMS treatment, when depressive symptoms start to decline. 49 patients with a diagnosis of major depressive disorder received open-label TMS treatment including 20 daily sessions of intermittent theta-burst stimulation (iTBS), a condensed TMS protocol [5]. We analysed how behavioural and neural markers of positive bias changed early on during treatment (after 8 iTBS sessions), and which of these changes were related to clinical outcome at the end of treatment.

Positive bias was assessed using a behavioural task (Facial Expression Recognition Task [19]) and a functional magnetic resonance imaging (fMRI) paradigm assessing implicit processing of emotional faces [20]. Both paradigms have been shown to capture negative biases in depression, and to be sensitive to modulation by antidepressant treatment [8, 20–24]. Positive bias in the FERT was defined as increased accuracy or speed in the recognition of positive compared to negative faces, and/or a general response bias towards indicating a positive emotion (rather than a negative one). In the fMRI paradigm, positive bias was defined as increased BOLD response towards happy compared to fearful faces. An increase in positive bias might be observed in task-positive networks, which show a positive blood-oxygen-level-dependent (BOLD) response to external stimuli [25], or task-negative networks (i.e. default mode network (DMN)) which show a negative BOLD response to external stimuli [26, 27].

Based on the evidence reviewed above, we hypothesised that an early increase in these behavioural and neural markers of positive bias would be related to improved clinical outcome at the end of treatment. In line with our pre-registered predictions, our findings demonstrate that clinical response at the end of treatment was associated with an early increase in response bias towards classifying facial expressions as positive (rather than negative). On a neural level, clinical improvement at the end of treatment was associated with an early increase in positive bias in the response of the rostral ACC to emotional faces. This change was accompanied by increased positive bias in task-related connectivity between rostral ACC and posterior DMN regions. These neural changes in positive bias predicted clinical improvement at the end of treatment beyond early symptom reduction. This study advances our understanding of mechanisms of action of TMS treatment and potential differences to antidepressant medication.

## 2 Results

### 2.1 Time course of HAM-D Improvement

49 patients diagnosed with major depressive disorder received open-label iTBS treatment including 20 daily sessions over the course of 4 weeks (Figure 1). The study included two experimental sessions, one before the start of treatment (‘Baseline’) and one after around 8 iTBS sessions (‘Week 2’), which included clinical questionnaires, performance of the Facial Expression Recognition Task (FERT) [19] and an fMRI paradigm assessing passive processing of emotional faces (gender discrimination task) [20]. The primary clinical outcome measure was the Hamilton Depression Rating Scale (HAM-D)[28] which was assessed at baseline and once per week during treatment, and at a two-week follow-up (Week 6). Demographic and clinical baseline characteristics are included in Table 1. 30 patients were on a stable dose of psychoactive medication (at least 6 weeks prior to the Baseline session)(Supplementary Table 1), the remaining patients were unmedicated.

**Figure 1.**
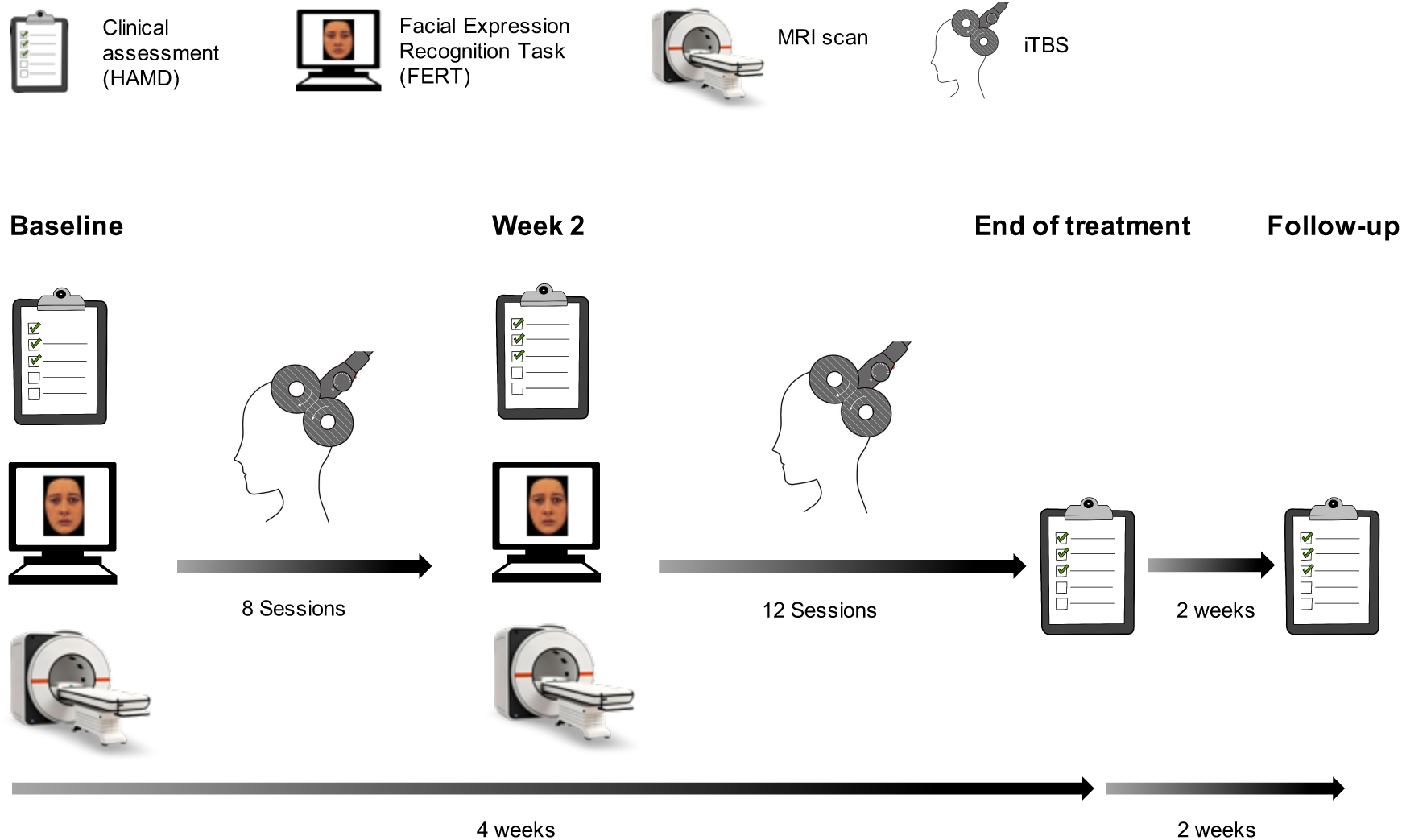
Experimental procedure. Participants completed a ‘Baseline’ experimental session involving clinical assessments, the Facial Expression Recognition Task, and an MRI scan (including task fMRI). After ∼8 iTBS sessions, a second experimental session (‘Week 2’) was conducted which was identical to the Baseline session. Participants received a total of 20 iTBS sessions over 4 weeks. Clinical assessments were completed at the end of treatment (‘Week 4’) and two weeks after the end of treatment (‘Week 6’). Figure adapted from [29].

**Table 1.** Baseline characteristics

|  | **Responders**<br>**(*n* = 33)** | **Non-Responders**<br>**(*n* = 16)** |
| --- | --- | --- |
| **Gender** |  |  |
| Female (%) | 28 (85%) | 13 (81%) |
| Male (%) | 5 (15%) | 3 (19%) |
| Age in years, mean (SD) | 39.9 (10.9) | 37.6 (9.2) |
| HAM-D, mean (SD) | 18.3 (2.8) | 17.6 (2.3) |
| MADRS, mean (SD) | 24.1 (4.3) | 26.0 (5.3) |
| YMRS, mean (SD) | 0.91 (1.1) | 0.88 (1.3) |
| **PANAS** |  |  |
| Positive, mean (SD) | 15.7 (3.9) | 14.9 (3.1) |
| Negative, mean (SD) | 29.2 (6.7) | 31.2 (7.5) |
| **STAI** |  |  |
| Trait, mean (SD) | 64.5 (7.4) | 61.0 (7.4) |
| State, mean (SD) | 66.9 (6.9) | 63.0 (8.0) |
| **Use of psychoactive medication** |  |  |
| No. taking medication (%) | 19 (58%) | 11 (69%) |

HAM-D: Hamilton Depression Rating Scale, MADRS: Montgomery-Asberg Depression Rating Scale [30], YMRS: Young Mania Rating Scale [31], PANAS: Positive and Negative Affect Schedule [32], STAI: and State-Trait Anxiety Inventory [33].

The time course of HAM-D scores from Baseline to Week 6 is displayed in Figure 2 (Supplementary Table S2). The average reduction in HAM-D score was 45.4% after around 8 iTBS sessions (Week 2), and 59.7% at the end of treatment (Week 4). 33 participants were classified as Responders, and 16 as Non-Responders (Week 4).

**Figure 2.**
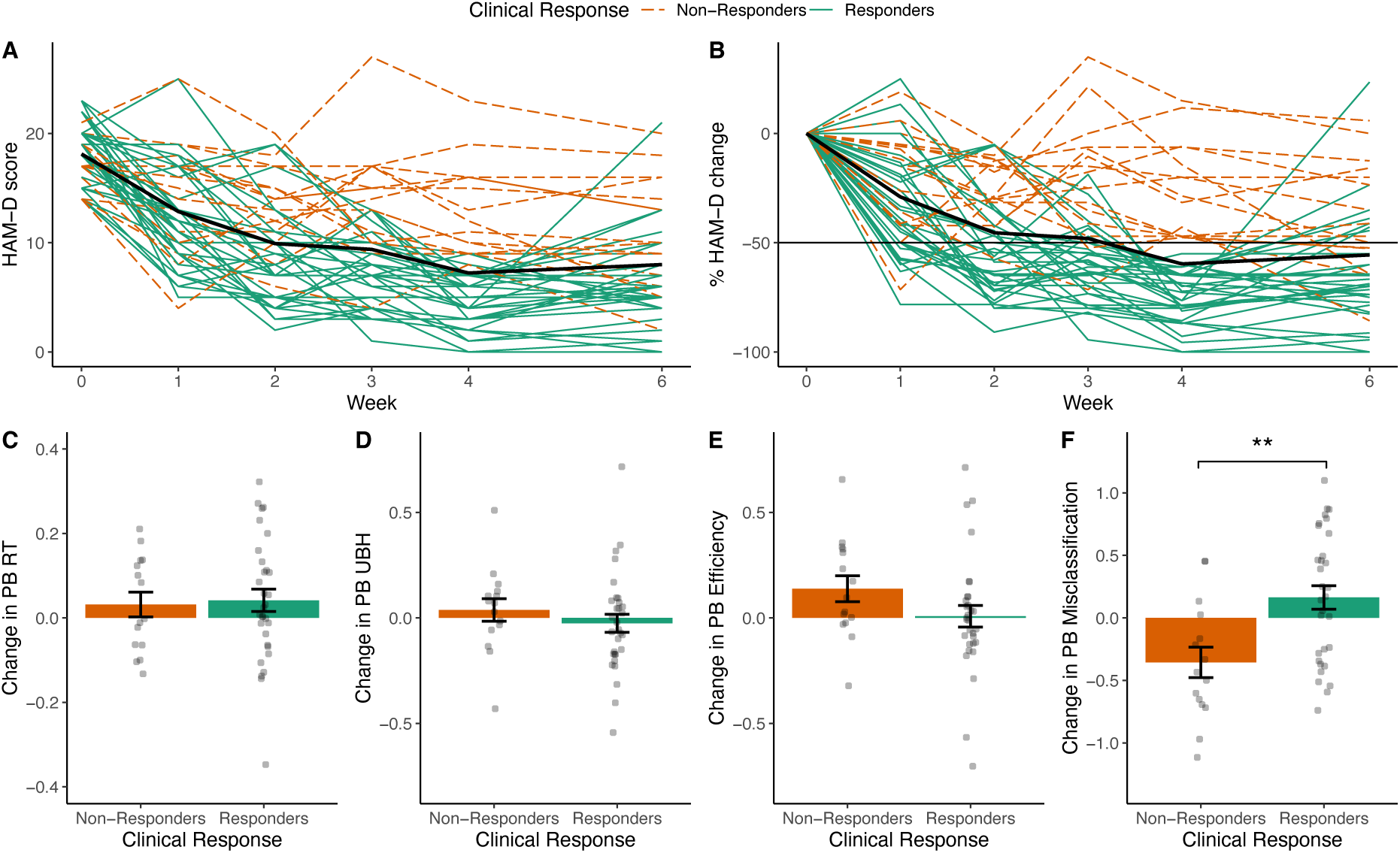
(A, B) Time course of HAM-D scores from Baseline (Week 0) to Week 6. (A) Raw HAM-D scores. (B) HAM-D scores as percentage change from Baseline (i.e. 100 * (HAM-D – Baseline HAM-D) / Baseline HAM-D). A negative value indicates a reduction in HAM-D score over time. Based on percentage reduction in HAM-D score from Baseline to Week 4, 33 participants were classified as Responders (≥50% reduction in HAM-D score), and 16 as Non-Responders. The thick black line represents the group average. (C-F) Change in positive bias measures from the FERT as a function of Clinical Response. Responders showed a relatively larger increase in Positive Bias Misclassification than Non-Responders (F). ** indicates .001 < *p* < .01. PB = Positive Bias, RT = Reaction Time, UBH = Unbiased Hit Rate. Error bars represent the standard error of the mean (SEM).

### 2.2 TMS induces positive response bias in categorisation of facial expressions

To analyse whether early changes in positive bias in the FERT were related to clinical outcome at the end of treatment, we assessed four positive bias measures which were defined based on different performance metrics (*Unbiased Hit Rate (UBH)*: classification accuracy, *Reaction Time*: response speed, *Efficiency*: Unbiased Hit Rate / Reaction Time, *Misclassifications*: frequency of an emotion chosen incorrectly, see Methods). Positive Bias Misclassification changed from Baseline to Week 2 as a function of Clinical Response (Clinical Response x Session interaction: *F*(1,44) = 10.5, *p* = .002, *p_corr_* = .009). Responders showed a relative increase in Positive Bias Misclassification compared to Non-Responders (*Cohen’s d* = 1.31, 95% CI = [0.85, 1.76]; Figure 2, Supplementary Figure S1, Supplementary Table S3), i.e. they misclassified more faces as positive (independent of the valence of the presented face). A logistic regression analysis controlling for Baseline HAM-D, Age, Gender, Psychoactive Medication and Number of iTBS Sessions (before the second experimental session in Week 2) confirmed that early change in Positive Bias Misclassification predicted Clinical Response at the end of treatment (Clinical Response: *β* = 2.18, *z* = 2.61, *p* = .009, *p_corr_* = .036, Odds Ratio = 3.39, 95% CI = [1.5, 9.7]). The change in the remaining positive bias measures was not related to Clinical Response (Positive Bias Reaction Time: Clinical Response x Session: *F*(1,44) = 0.05 *p* = .82; Positive Bias UBH: *F*(1,44) = 0.77, *p* = .39; Positive Bias Efficiency: *F*(1,44) = 2.3, *p* = .13). The changes in positive bias measures did not predict HAM-D Improvement as continuous measure (Supplementary Figure S2).

A follow-up ANOVA was run to test whether the relative increase in Positive Bias Misclassification in Responders was driven by a change in a particular emotion. There was a significant interaction effect between Clinical Response, Session and Emotion (*F*(5,220) = 2.4, *p* = .045). Post-hoc ANOVAs for individual emotions did not show any significant interactions between Clinical Response and Session (all *p_corr_* > .25), suggesting that the change in Positive Bias Misclassification was based on a change in the overall pattern in the data rather than a specific emotion (Supplementary Figure S1).

### 2.3 TMS induces positive bias in neural response to emotional faces

The fMRI paradigm presented happy faces and fearful faces in a block design. Participants were asked to indicate the gender of each face. In line with previous studies, presentation of happy and fearful faces (Happy + Fearful contrast) elicited a positive BOLD response in large areas across the brain, including ACC, insula and amygdala (Figure 3A). As expected, areas associated with the default mode network (such as precuneus and medial prefrontal cortex) showed a negative BOLD response (- (Happy + Fearful) contrast)(Figure 3B, Supplementary Table S4).

**Figure 3.**
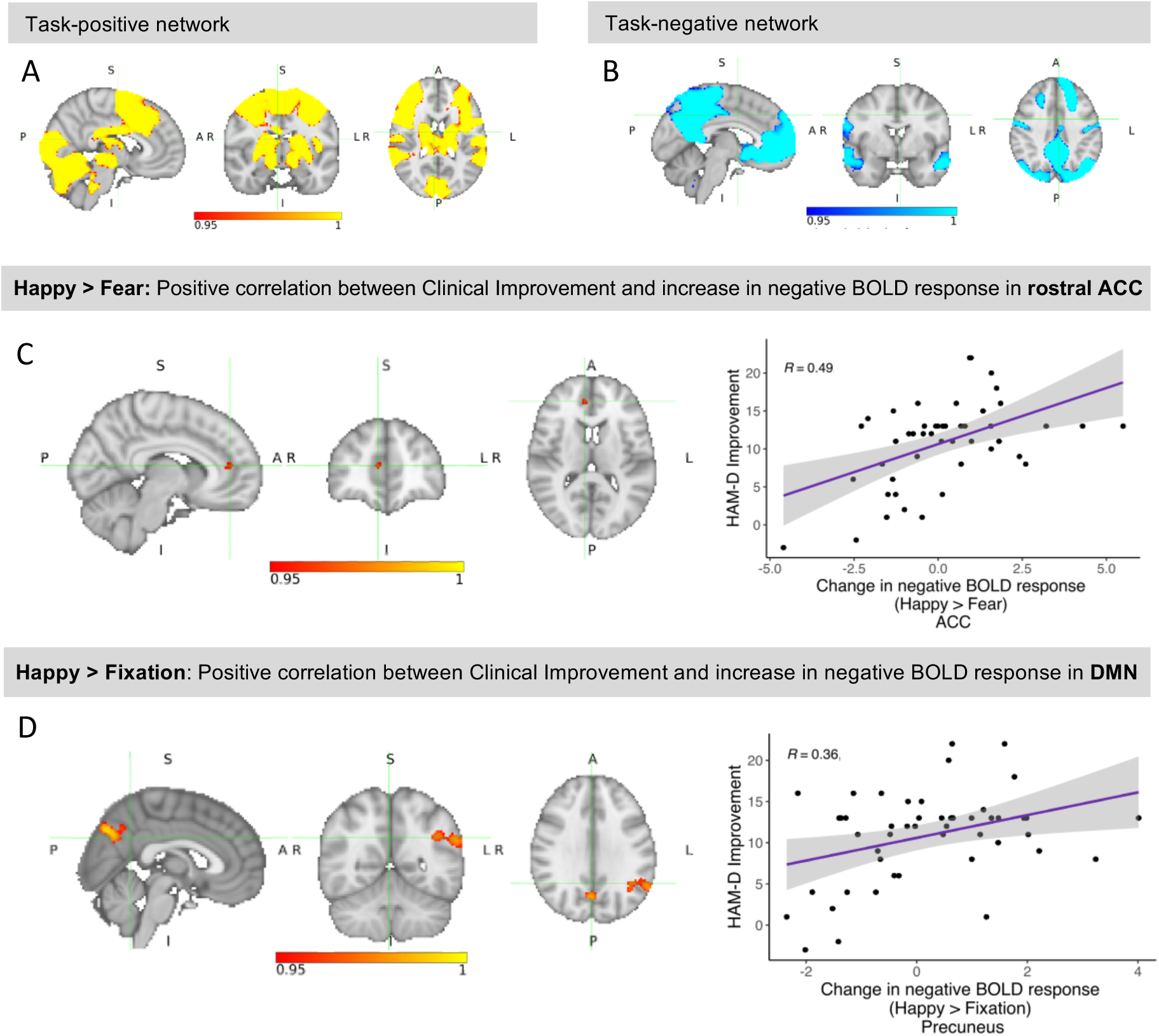
Group-average task activation: (A) Task-positive network (mean BOLD response to happy and fearful faces). The task activated a single large cluster spanning various areas across the brain (cluster size: 82,062 voxels). The maximum was located in the left temporal occipital fusiform cortex (*tmax* = 16.5, x = -38, y = -50, z = -22, *p* < .001). (B) Task-negative network (negative mean response to happy and fearful faces). Seven clusters showed a negative BOLD response during task performance, including medial prefrontal cortex, rostral ACC, precuneus, posterior cingulate and angular gyrus. The maximum was located in the left lateral occipital cortex (*tmax* = 13.9, x = -40, y = -70, z = 34, *p* < .001). (C) In the Happy > Fearful contrast (whole-brain), HAM-D Improvement correlated positively with an increase in negative BOLD signal in a single cluster in the rostral ACC which overlapped with the task-negative network shown in (B)(17 voxels, *tmax* = 5.35, x = 8, y = 44, z =12, *p* = .045). (D) In the Happy > Fixation contrast, HAM-D Improvement correlated with an increase in negative BOLD response in 3 clusters overlapping with the task-negative network, including left precuneus, angular gyrus, and supramarginal gyrus (maximum in the precuneus: *tmax* = 5.70, x = 2, y = -74, z = 38, *p* = .008). Please note that a positive value for ‘change in negative BOLD response’ indicates an increase in negative BOLD response (i.e. more negative BOLD signal).

In the whole-brain analysis, HAM-D Improvement correlated positively with an increase in negative BOLD response (i.e. increase in de-activation, see [34]) in the Happy > Fearful contrast in the rostral ACC, i.e. with an increase in positive bias in rostral ACC response (17 voxels, *tmax* = 5.35, x = 8, y = 44, z =12, *p* = .045) (Figure 3C). Visualisations of the correlation suggest that this effect was mainly driven by a positive correlation between HAM-D Improvement and increase in negative BOLD response to happy faces (Supplementary Figure S3). ROI analysis did not reveal any further correlations with HAM-D Improvement.

We further analysed the constituent contrasts Happy > Fixation and Fearful > Fixation. In the whole-brain analysis, HAM-D Improvement correlated with an increase in negative BOLD response for Happy > Fixation in three clusters overlapping with the task-negative network, including left precuneus, angular gyrus, and supramarginal gyrus (maximum in the precuneus: *tmax* = 5.70, x = 2, y = -74, z = 38, *p* = .008) (Figure 3D, Supplementary Figure S4, Supplementary Table S5). This is in line with the findings above and suggests that the increase in positive bias might extend beyond the rostral ACC to other areas of the DMN. No significant clusters emerged in the Fearful > Fixation contrast, or in the ROI analysis.

### 2.4 TMS induces positive bias in task-related connectivity

To further investigate our main finding, the correlation between HAM-D Improvement and increase in negative BOLD response for Happy > Fearful faces in the rostral ACC, we conducted a psycho-physiological interaction (PPI) analysis [35]. In contrast to the analyses described above which focused on brain *activation*, PPI analysis captures task-specific changes in the *functional connectivity* between different brain areas. PPI analysis identifies brain areas which show an increase in connectivity (i.e. enhanced communication) with a selected seed region in a task-specific psychological context. We ran a whole-brain PPI analysis based on the Happy > Fearful contrast using the significant cluster in the rostral ACC as seed region. Significant clusters in the Happy > Fearful PPI contrast represent regions which showed a stronger correlation with the rostral ACC seed region during the presentation of happy compared to fearful faces (i.e. positive bias in connectivity).

The PPI analysis revealed a positive correlation between HAM-D Improvement and increase in PPI in the Happy > Fearful contrast in several regions of the DMN (precuneus, posterior cingulate), sensorimotor areas (precentral and postcentral gyrus) and visual areas (occipital lobe)(Figure 4, Supplementary Figure S6, Supplementary Table S6). This indicates that HAM-D Improvement was associated with a relative increase in connectivity of rostral ACC with the identified areas in response to happy faces (compared to fearful faces), i.e. increased positive bias in connectivity.

**Figure 4.**
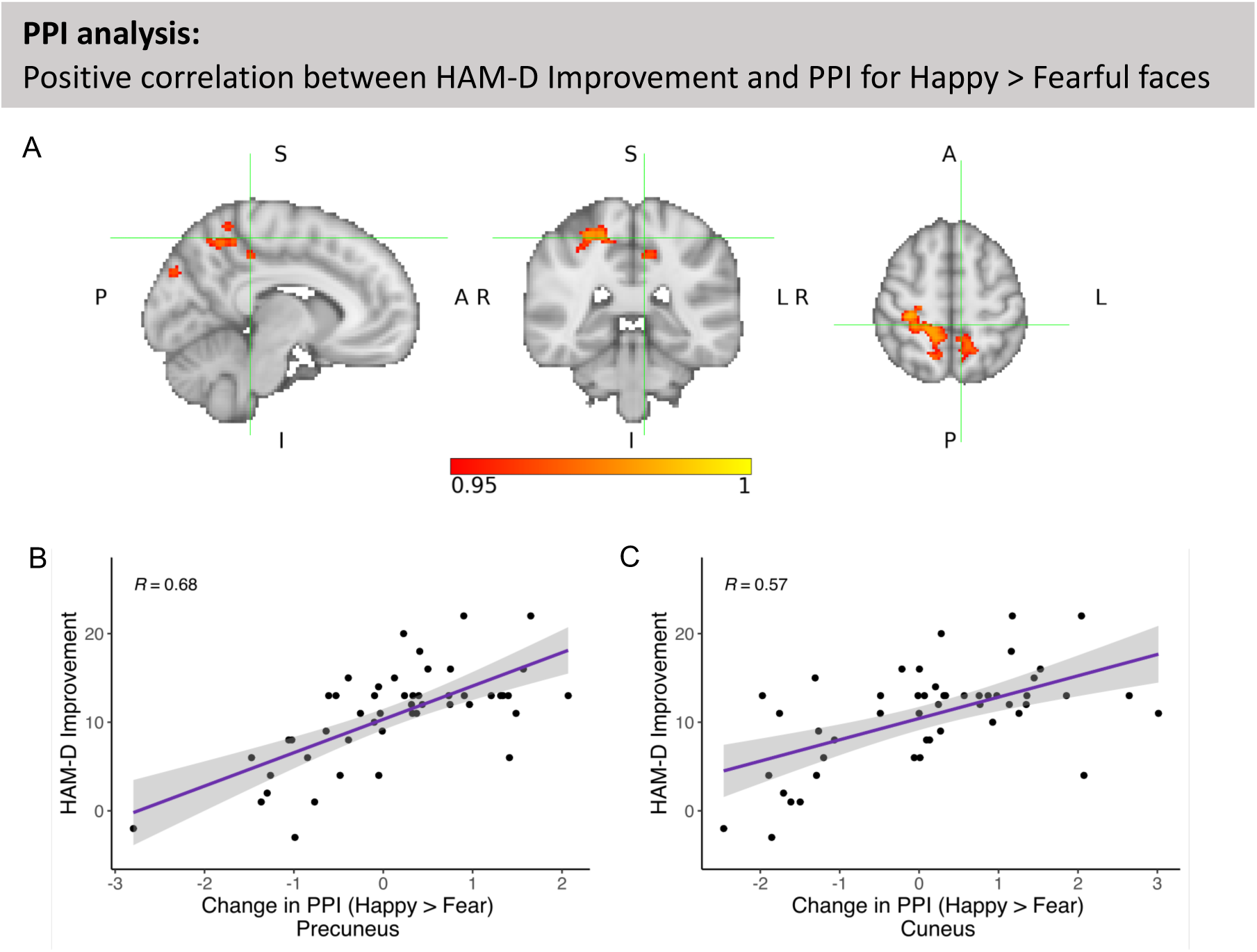
(A) A psycho-physiological interaction (PPI) analysis with rostral ACC as seed region revealed a positive correlation between HAM-D Improvement and change in PPI in the Happy > Fearful contrast in various areas of the DMN, sensorimotor and visual areas. The peak voxel was located in the right precentral gyrus (*tmax* = 4.78, x = 26, z = -24, z = 60, *p* = .014). This suggests that individuals with larger HAM-D Improvement showed a relative increase in connectivity between those areas and rostral ACC when happy faces were presented, and a relative decrease in connectivity when fearful faces were presented, i.e. an increase in positive bias in connectivity. (B) and (C) show the correlation between HAM-D Improvement and increase in PPI for happy vs. fearful faces in the two largest clusters spanning the left and right precuneus, precentral gyrus, left posterior cingulate, and right postcentral gyrus (B), as well as left cuneus and lateral occipital cortex (C).

### 2.5 TMS induces positive bias in the fMRI gender discrimination task

While the primary objective of the fMRI paradigm was to assess neural processing of emotional information, we also analysed performance in the gender discrimination task. Regression analysis suggests that an increase in reaction time (RT) for happy faces was a significant predictor of HAM-D Improvement, such that a larger increase in RT from Baseline to Session 2 predicted larger HAM-D Improvement (*β* = 0.015, *t*(35) = 2.03, *p* = .0491)(Figure S8). The change in RT for fearful faces was not related to HAM-D Improvement (Fearful: *β* = 0.011, *t*(35) = 1.33, *p* = .19). Response accuracy in the gender discrimination task was not related to Clinical Response or HAM-D Improvement (all *p* > .16, Figure S8).

An increase in RT for happy faces can be interpreted as attentional capture, i.e. the happy facial expressions might distract participants from the task (which is gender discrimination), which would be in line with an increase in positive bias. To test whether the increase in RT for happy faces was related to the increase in positive bias in brain activity and connectivity observed in the fMRI analysis, we calculated correlations between the change in RT for happy faces with the changes in brain activity and connectivity reported above. The increase in RT for happy faces correlated positively with the increase in response to happy faces in the precuneus (*r* = 0.4, *t*(40) = 2.7, *p* = .0083), and with the increase in connectivity (PPI) for happy vs. fearful faces in the precuneus (*r* = 0.41, *t*(40) = 2.8, *p* = .0073) and cuneus (*r* = 0.5, *t*(40) = 3.6, *p* = .0007) (Figure S9).

### 2.6 Behavioural and neural increases in positive bias predict clinical response independently

Our analyses above revealed two main findings: an early increase in positive response bias in the FERT (Positive Bias Misclassification) predicted Clinical Response at the end of treatment, and an early increase in activation to positive compared to negative faces in the rostral ACC was associated with HAM-D Improvement at the end of treatment. To test whether early changes in these behavioural and neural markers of positive bias predict Clinical Response independently, we ran a logistic regression analysis including the change in the behavioural and neural markers as regressors of interest. Both regressors were significant, indicating that the behavioural and neural changes were independent predictors of Clinical Response (change in rostral ACC: *β* = 1.13, *z* = 2.74, *p* = .0061, Odds Ratio = 8.25, 95% CI = [2.31, 53.51]; change in Positive Bias Misclassification: *β* = 2.8, *z* = 2.42, *p* = .015, Odds Ratio = 4.84, 95% CI = [1.61, 22.33]).

### 2.7 Neural changes predict HAM-D Improvement beyond early symptom reduction

By the second testing time point (Week 2), HAM-D scores had already substantially improved (Figure 2). This raises the question of whether the changes in neural and behavioural measures of positive bias purely reflected early symptom improvement, or whether they predict HAM-D Improvement at the end of treatment beyond the early change in HAM-D scores (from Baseline to Week 2). Regression analyses were run to predict HAM-D Improvement at the end of treatment including the early change in HAM-D score and the change in positive bias measures as regressors of interest (and Baseline HAM-D, Age, Gender, Psychoactive Medication, and Number of TMS Sessions as control regressors). Separate regression analyses were run for each positive bias measure found to be related to clinical outcome in the analyses above (change in rostral ACC response (Happy > Fear), precuneus and angular gyrus response (Happy > Fixation), PPI in precuneus and cuneus (Happy > Fear), reaction time for happy faces (fMRI task), and in Positive Bias Misclassification (FERT)). All neural measures predicted Clinical Improvement beyond early HAM-D change (all p < .003, Tables S7-13). The strongest predictor was the change in PPI in the precuneus (*β* = 2.8, *t*(41) = 4.5, *p* < .0001, *partial R^2^* = 0.33, Table S10). The behavioural measures did not significantly predict clinical outcome beyond the early clinical change (see Supplementary Material).

These findings suggest that the neural changes in positive bias might predict HAM-D Improvement beyond early HAM-D change. To test whether early changes in neural markers might improve prediction of HAM-D Improvement in unseen data, we created machine-learning models (elastic net regression) which we trained and tested in a robust nested cross-validation procedure. An elastic net regression model trained on early HAM-D change (Week 2), Baseline HAM-D, Age, Gender, Psychoactive Medication achieved an *R^2^* value of 0.19 (SD = 0.24). Adding early changes in neural and behavioural markers of positive bias as predictors improved *R^2^* value to 0.52 (SD = 0.22)(Figure S11). Inspection of the regression weights confirmed that the improved prediction resulted from the neural markers rather than the behavioural ones (which were assigned a regression weight of zero)(Figure S12).

## 3 Discussion

The goal of this open-label iTBS treatment study was to test for the first time whether predictions derived from the neuropsychological theory of antidepressant action [7] generalise to TMS depression treatment. We hypothesised that early changes in behavioural and neural markers of positive bias would relate to clinical outcome at the end of treatment. With respect to behaviour, we found that clinical response was related to an increase in positive bias in misclassification of emotional faces. On the neural level, HAM-D improvement was associated with increased positive bias in negative BOLD response to emotional faces in the DMN including rostral ACC. Moreover, clinical improvement was related to an increase in positive bias in connectivity between rostral ACC and large areas across the brain including posterior DMN regions. These neural changes were correlated to increased positive bias in reaction times in the fMRI task. The increased positive response bias in emotion classification and neural changes in positive bias were independent predictors of clinical response at the end of treatment. The early change in neural measures predicted clinical improvement at the end of treatment beyond early clinical improvement.

Negatively biased information processing is theorised to play a causal role in the development and maintenance of depression [10]. In our study, we used the Facial Expression Recognition Task to assess biases towards positive or negative facial expressions. We found that positive bias in the misclassification of facial expressions changed as a function of clinical response. Patients who responded to iTBS treatment showed a relative increase in positive bias, which supports our hypothesis. Increased positive bias in misclassifications can be interpreted as an increased response bias towards positive emotions, i.e. tendency to interpret ambiguous facial expressions as positive (even if they are not). Similar increases in the processing of positive vs. negative information have been reported in response to other antidepressant treatments and might contribute to symptom reduction by disrupting negative thought patterns [10, 21, 22, 24].

Research on neural mechanisms of TMS treatment has mostly focused on resting-state connectivity. However, resting-state fMRI is unable to capture how the brain processes different types of stimuli in the environment, such as positive compared to negative stimuli. Task fMRI is able to capture such processes, is considered as being more interpretable, and might show stronger links to behaviour [36].

Using an fMRI task involving the presentation of happy vs. fearful faces, we found strong evidence for our hypothesis that an early increase in positive bias was related to HAM-D improvement. HAM-D improvement correlated with an increase in positive bias in negative BOLD response in the rostral ACC. Pre-treatment activity in the rostral ACC has been shown to predict treatment response to different types of antidepressant treatments [37], and response to TMS treatment has been associated with an increase in rostral ACC volume [38]. The rostral ACC is part of the default mode network which typically de-activates during the performance of cognitive tasks. Depression has been associated with reduced negative BOLD response of the DMN to external stimuli [34, 39], especially positive stimuli [40], which is hypothesised to cause rumination and reduced engagement with external stimuli. Our results suggest that TMS might normalise this deficit early on during treatment, by increasing de-activation of the DMN in response to positive external stimuli. We observed a similar relationship between clinical improvement and increase in positive bias in the precuneus and angular gyrus, suggesting that the positive bias induced by TMS extended beyond the rostral ACC to more posterior parts of the DMN. The increase in response to happy faces in the precuneus was correlated to an increase in reaction time to happy faces, showing that the neural change was underpinned by an increase in positive bias in behaviour (increased attentional capture by positive facial expressions).

Moreover, our PPI analysis demonstrates that TMS treatment did not only induce a positive bias in brain *activity*, but also in *functional connectivity* (i.e. communication between different brain areas). HAM-D improvement was associated with increased positive bias in connectivity between the seed region in the rostral ACC and posterior parts of the DMN (precuneus, posterior cingulate), sensorimotor and visual areas. In other words, patients who experienced larger clinical improvement showed an early increase in connectivity (i.e. communication) between rostral ACC and several cortical areas during the presentation of positive (compared to negative) stimuli. Depression has been associated with reduced connectivity of rostral ACC to posterior parts of the DMN (such as precuneus and posterior cingulate)[41], and sensorimotor regions [42]. Our findings suggest that iTBS treatment might normalise such impairments, which might lead to increased processing of positive information and enhanced emotional processing [41, 43]. In line with this, the changes in connectivity were correlated to increased positive bias in reaction times in the gender discrimination task, suggesting that the neural effects of TMS were strong enough to translate into changes in behaviour.

The aim of our study was to investigate potential *mechanisms* of TMS treatment. By the second testing time point, HAM-D scores had already substantially improved, so it is unclear whether the behavioural and neural changes in positive bias preceded clinical improvement. To rule out the possibility that the changes in positive bias might purely be a reflection of early symptom improvement, we conducted regression analyses to test whether changes in positive bias predicted clinical improvement at the end of treatment beyond the early clinical change (change in HAM-D score in Week 2). All neural measures predicted clinical improvement at the end of treatment beyond the early clinical change. Elastic net regression applied in a robust nested cross-validation framework confirmed that features based on neural markers of positive bias increased the percentage of explained variance in unseen data from 19% to 52%. This demonstrates that the neural changes were strongly related to clinical improvement even after accounting for the early clinical change, making it unlikely that the neural changes were purely a reflection of the early clinical change.

Taken together, our results provide strong evidence suggesting that the neuropsychological theory of antidepressant action generalises to TMS treatment. Multiple studies found that antidepressant drugs induced a positive bias in regions of the salience network such as dorsal ACC, insula and amygdala [8, 44–46]. Interestingly, we found no such effect in our study. Our results suggest that, like antidepressant drugs, TMS treatment might increase the processing of positive vs. negative information early on during treatment, but the neural mechanisms might differ. While drugs might primarily affect the salience network, TMS might primarily affect the DMN, likely via connections between DLPFC and subgenual ACC [13, 47–49].

This study has several limitations. First, the study did not include a control group (e.g. sham stimulation). It is therefore possible that the observed effects might in part be due to placebo effects rather than iTBS treatment. Second, symptoms had already improved substantially within 8 treatment sessions. Therefore, it is unclear whether changes in emotional processing preceded clinical improvement. Third, a large proportion of patients in our sample were taking a stable dose of psychoactive medication (which was controlled for in the analysis). Consequently, it cannot be ruled out that the effects observed in this study might partially be related to interactions with medication. Replication studies are necessary to assess the replicability and generalisability of our findings.

## 4 Methods

This study has been pre-registered (NCT04969549, https://clinicaltrials.gov/study/NCT04969549 and https://osf.io/rhxm2).

### 4.1 Sample

The **Bra**zil-**En**gland Study on **M**echanisms **a**nd Response **P**redictors of iTBS Treatment (BRAEN-MAP study) was conducted in the Instituto de Psiquiatria do Hospital das Clínicas da Faculdade de Medicina da Universidade de São Paulo between August 2021 and November 2022. Participants were recruited through social media advertisement, Google Ads and flyers posted on bulletin boards at the Hospital das Clínicas. 52 individuals took part in the study. 3 participants discontinued the study after two weeks of iTBS treatment, resulting in a final sample size of 49 participants (41 female). Participants were eligible for the study if they were between 18 and 65 years of age, had a Hamilton Depression Rating Scale (HAM-D)[28] score of 14 or above, and had a current diagnosis of major depressive disorder as confirmed by the Mini-International Neuropsychiatric Interview (MINI)[50]. Exclusion criteria were current diagnosis of substance dependence (such as alcohol), severe clinical or neurological disorders, suicidal ideation, presence of psychotic symptoms, severe depression characterised by a HAM-D score greater than 28, manic symptoms as indicated by a score above 8 in the Young Mania Rating Scale. Patients with contraindications to TMS or MRI, such as metallic implants or a diagnosis of epilepsy, were excluded from the study. 30 of the participants were taking psychoactive medication (see Table S1). Clinical scores at Baseline are included in Table 1. The study was approved by the National Ethics Committee in Brazil (CAEE: 52384921.6.0000.0068).

### 4.2 Procedure

Participants were invited to a ‘Baseline’ experimental session which included clinical assessments, a behavioural task (Facial Expression Recognition Task (FERT)), and an MRI scan, including a structural T1 scan and task fMRI (other modalities were acquired but are not part of this report)(Figure 1). After completion of the Baseline session, participants started iTBS treatment which consisted of 20 sessions of iTBS delivered over the course of four weeks (Monday to Friday). The protocol included 1,800 pulses per session applied at 100% resting motor threshold. The resting motor threshold was defined as the lowest intensity required to elicit a contraction in the first dorsal interosseous muscle in 3 out of 5 TMS pulses over the motor hotspot. The resting motor threshold was assessed on the first two days of each treatment week. If there was a significant change of more than 5 percentage points between the two days, the motor threshold was reassessed on the third day (or even further), until there was no significant variation from one day to the next.

After around 8 iTBS sessions, participants completed a second experimental session (‘Week 2’) including the same clinical assessments, behavioural task, and MRI as in the ‘Baseline’ session. Due to practical considerations, the exact number of iTBS sessions varied between 7 and 10. Most participants completed the second experimental session after 8 iTBS sessions (4/37/4/4 participants received 7/8/9/10 iTBS sessions before the second experimental session). Participants completed the clinical assessments again at the end of treatment (Week 4), and two weeks after the treatment completion as follow-up (Week 6).

### 4.3 Clinical and questionnaire measures

The primary outcome measure was the change in Hamilton Depression Rating Scale (17-item version)(HAM-D)[28] score from Baseline to the end of treatment (Week 4). *Clinical Response* was defined as a categorical variable reflecting a reduction in HAM-D score from Baseline to Week 4 by at least 50% (‘Responders’), or less than 50% (‘Non-Responders’). Since the use of a continuous outcome variable might increase statistical power [51], *HAM-D Improvement* was used as additional continuous outcome variable, defined as the improvement in HAM-D score from Baseline to Week 4 (i.e. Baseline HAM-D – Week 4 HAM-D, a positive value indicates symptom reduction). Additional clinical assessments are included in Table 1.

The HAM-D score at Week 4 was missing for one participant (Week 6 score was available). Reduction in HAM-D score at Week 4 was estimated by fitting an exponential decay function to the remaining HAM-D reduction scores for this participant (Week 0-3).

### 4.4 Facial Expression Recognition Task (FERT)

The FERT was adapted from [19]. The task protocol included 250 trials in total, separated into two blocks of 62 and two blocks of 63 trials. On each trial, a face with an emotional expression was presented for 500ms. Participants were instructed to indicate the emotion of the face as quickly and accurately as possible by clicking onto the corresponding key on a labelled keyboard. Emotions included happy, surprise, anger, fear, sadness, disgust and neutral. Each emotion (apart from neutral) was presented at 10 different intensities, by four different actors (i.e. 4 x 10 images per emotion). Ten neutral face were included as well.

As outcome measures, four Positive Bias measures were defined based on different performance metrics. These performance metrics were calculated separately for positive (happiness, surprise) and negative emotions (fear, anger, sadness, disgust). Positive Bias measures were defined as the ratio of each performance metric for positive vs. negative stimuli, and were log-transformed to ensure symmetry and interpretability. All Positive Bias measures were defined in a way so that a higher value indicates a more positive bias.

#### Positive Bias Unbiased Hit Rate

The Unbiased Hit Rate (based on signal-detection theory [52]) is a measure for how accurately participants identified each emotion. It takes into account how often each emotion was correctly identified, but also how often each emotion was chosen incorrectly:

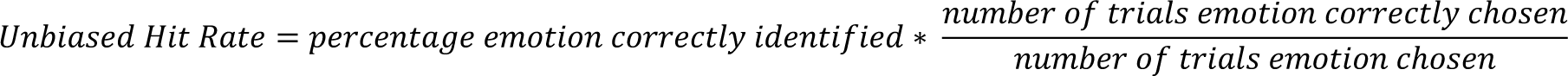

The measure of interest was *Positive Bias Unbiased Hit Rate*, defined as the log-transformed ratio of Unbiased Hit Rate averaged across positive emotions (happy, surprise) divided by averaged Unbiased Hit Rate for negative emotions (fear, sadness, anger, disgust).

#### Positive Bias Reaction Time

For each emotion, the median reaction time was calculated by taking the median across all trials in which the emotion was correctly identified. *Positive Bias Reaction Time* was calculated as the log-transformed ratio of the median reaction time for negative emotions divided by the median reaction time for positive emotions.

#### Positive Bias Efficiency

*Efficiency* combines accuracy and speed into one measure, by dividing *Unbiased Hit Rate* by the median reaction time. Positive Bias Efficiency was defined as the log-transformed ratio of *Efficiency* for positive emotions divided by *Efficiency* for negative emotions.

#### Positive Bias Misclassification

In addition to the measures specified in our preregistration, we also analysed misclassifications. The number of Misclassifications for each emotion was the number of times the emotion has been chosen incorrectly. *Positive Bias Misclassification* was defined as the log-transformed ratio of Misclassifications for positive emotions, divided by the number of Misclassifications for negative emotions (this is independent of whether the correct emotion was positive or negative). In contrast to the above measures which capture a bias in the ability to recognise emotions, this measure captures a bias in the response criterion (i.e. how often participants indicated that a positive (vs. negative) emotion was shown, although a different emotion was actually presented), and has been found to be sensitive to the effects of antidepressant medication [22].

### 4.5 FERT analysis

FERT data were analysed in R (version 4.1.1) [53]. To test whether changes in positive bias were related to Clinical Response, an ANOVA was conducted for each positive bias measure as dependent variable, Time (Baseline vs. Week 2) as within-subject factor and Clinical Response (Responders vs. Non-Responders) as between-subject factor. Significant findings were cross-validated using a logistic regression analysis predicting Clinical Response. Change in positive bias was included as regressor of interest, and Baseline HAM-D, Age, Gender, use of Psychoactive Medication (yes/no) and Number of iTBS Sessions (before ‘Week 2’ time point) as control regressors. Significant interaction effects of Clinical Response and Time were followed up using an ANOVA to test whether the effect was driven by a specific emotion (change in positive bias as dependent variable, Emotion (Happiness, Surprise, Fear, Anger, Sadness, Disgust) and Time as within-subject factors, and Clinical Response as between-subject factor).

To test whether changes in positive bias might predict HAM-D Improvement as continuous variable, linear regression analyses were performed separately for each positive bias measure, using HAM-D Improvement as dependent variable, and change in positive bias as predictor. Baseline HAM-D, Age, Gender, Psychoactive Medication and Number of iTBS Sessions were included as control regressors.

Reported p-values are either uncorrected (‘*p*’), or corrected for multiple comparisons using Bonferroni correction (‘*p_corr_’*) (analysis of positive bias measures was corrected for the number of positive bias measures (four), analysis of individual emotions was corrected for the number of emotions (six)).

### 4.6 Emotional Faces fMRI Task

Participants completed a gender discrimination task which involved passive processing of emotional facial expressions [20]. The task included four blocks of happy faces and four blocks of fearful faces, which were interleaved by nine blocks displaying only a fixation cross. In total, 120 fearful and 120 happy faces were presented for 100ms each. Participants were instructed to report the gender of the presented face via button press. Although the emotional expression of the faces is irrelevant to the task, the task has been shown to activate emotional processing networks [8, 20]. We did not expect TMS treatment to have an effect on performance in the gender discrimination task, but have included an analysis of task performance in the Supplementary Material for completeness (Supplementary Figure S3).

### 4.7 FMRI pre-processing

FMRI data were analysed using FSL (FMRIB Software Library)[54]. Structural T1-weighted images were brain-extracted using the Brain Extraction Tool (BET)[55]. Motion correction using MCFLIRT was applied [56, 57]. FMRI data were smoothed using a Gaussian kernel with Full Width Half Maximum of 6mm, and normalised using grand mean scaling. A high-pass filter with a cut-off of 100s was used. To correct for distortions, B0 unwarping was applied using field map images (rads and magnitude). Since some field maps included distortions, affected field maps were replaced by a mean field map calculated across 20 undistorted field map images [58]. For the first-level analysis, FILM pre-whitening was applied.

For advanced artefact removal, the task fMRI data were denoised using independent component analysis. The MELODIC (multivariate exploratory linear optimized decomposition into independent components) tool was used to decompose the data into independent components [59]. 20 randomly chosen datasets were manually labelled as signal, noise or unknown [60]. FIX (FMRIB’s ICA-based Xnoiseifier)[61, 62] was trained on the hand-labelled data, and the trained algorithm was then applied to all datasets. Components labelled as noise were removed from the signal, whereas components labelled as signal or unknown were retained.

FMRI images were registered to the high-resolution T1-weighted images via Boundary-Based Registration (BBR) using FMRIB’s Linear Image Registration Tool (FLIRT) [56, 57]. Non-linear transformations from structural to MNI space were calculated using FMRIB’s Non-Linear Image Registration Tool (FNIRT) [63]. These non-linear transformations were applied to the contrast images for higher-level analyses.

### 4.8 FMRI Analysis

FMRI data were analysed using FSL (FMRIB Software Library)[54]. Data and task design were modelled using General Linear Models (GLM) in FEAT (FMRI Expert Analysis Tool) by convolving a gamma hemodynamic response function with the task regressors for the presentation of happy and fearful faces. The temporal derivatives were added as additional regressors. On the first level, the main contrast of interest was Happy > Fearful, but the constituent contrasts Happy > Fixation and Fearful > Fixation were analysed as well. As additional method for motion correction, each individual’s motion parameter estimates from MCFLIRT were included in the GLM (6 additional regressors for rotation or translation). On the second level, a fixed-effects analysis was run for each individual, to assess increase in BOLD response from Baseline to Week 2 (Week 2 > Baseline).

On the third level, data from all participants were combined in a mixed-effects analysis to test for relationships between increase in positive bias from Baseline to Week 2 and clinical outcome at Week 4. Changes in positive bias might be observed in task-positive networks (positive BOLD response) or task-negative networks (negative BOLD response). We refer to increases in de-activations (i.e. more negative BOLD signal) in task-negative networks as ‘increase in negative BOLD response’(this terminology was adopted from [34]). Please note that an increase in positive bias can also be interpreted as a change in negative bias in the opposite direction (e.g. an increase in Happy > Fear is equivalent to a decrease in Fear > Happy). Statistical inference was performed using Randomise (FSL’s tool for nonparametric inference) [64] and threshold-free cluster enhancement (TFCE) [65] controlling the family-wise error rate at 0.05. The GLM included HAM-D Improvement as continuous regressor to test for correlations with changes in brain activity.

Additional analyses were run including Clinical Response as categorical regressor (instead of HAM-D Improvement), which yielded similar results (see Supplementary Material and Supplementary Figure S5). All analyses included Baseline HAM-D, Age, Gender, Psychoactive Medication and Number of iTBS Sessions (before the second fMRI scan) as control regressors of no interest. All regressors were de-meaned.

In addition to whole-brain analyses, region-of-interest (ROI) analyses were performed for our a priori ROIs (left and right amygdala, left and right insula, ACC, ventral striatum). Small volume correction analyses were performed using anatomical masks based on the Harvard-Oxford Structural Atlas in FSL (threshold = 50%, i.e. all voxels with a probability of belonging to the ROI of at least 50%) based on the GLMs described above. The mask for the ventral striatum was derived from the Oxford-GSK-Imanova Structural–anatomical Striatal Atlas. Significance within the anatomical masks was assessed using TFCE and Randomise. To correct for multiple comparisons across the six ROIs, the alpha level was set to 0.05/6 = 0.0083 (maps thresholded at 1-*p* = 0.9917).

To visualise the results, individual parameter estimates for significant clusters were extracted from the first-level statistical maps (using Featquery).

To further investigate our main finding, a correlation between Clinical Improvement and increase in negative BOLD response for Happy > Fear in the rostral ACC, we conducted a psycho-physiological interaction (PPI) analysis [35] using the significant cluster in the rostral ACC as seed region. A binary mask was created based on the significant cluster in the rostral ACC. This mask was warped into individual participant space. The *Fslmeants* tool was used to extract the timecourse of the signal within the mask for each participant and each session. The design matrix on the first level included regressors for the presentation of fearful and happy faces, one regressor representing the mean timecourse extracted from the rostral ACC mask, a PPI regressor for happy faces (interaction between Happy regressor and timecourse extracted from the rostral ACC mask), and a PPI regressor for fearful faces (interaction between Fearful regressor and timecourse extracted from the rostral ACC mask). The contrast of interest was Happy > Fearful, i.e. brain regions that show higher connectivity with rostral ACC during the presentation of happy compared to fearful faces (i.e. positive bias in connectivity). In line with the task fMRI analysis described above, the second level combined Baseline and Week 2 sessions for each participant to calculate the change between them (Week 2 > Baseline). On the third-level, group-level statistics were evaluated using Clinical Improvement as regressor of interest. The analysis included Baseline HAM-D, Age, Gender, Psychoactive Medication and Number of iTBS Sessions (before the second fMRI scan) as control regressors of no interest.

To ensure that our results were not confounded by changes in motion between sessions or groups, we analysed whether estimates of absolute or relative motion changed over time and whether there was a relationship with clinical outcome. There was no evidence for such potential confounds (see Supplementary Material, Figure S10).

### 4.9 Analysis of task performance in the fMRI gender discrimination task

For each participant, the median reaction time (RT) was calculated separately for happy faces and fearful faces (correct responses only). Accuracy was defined as the percentage of correct responses for happy or fearful faces. RT and accuracy were analysed in repeated-measures ANOVAs, including Session, Emotion and Clinical Response as independent variables (see Supplementary Material).

### 4.10 Combined behavioural and fMRI analysis

To test whether the observed behavioural (change in Positive Bias Misclassification) and neural (change in rostral ACC in Happy > Fearful contrast) changes in positive bias are independent predictors of the clinical outcome at the end of the treatment, we ran a logistic regression analysis to predict Clinical Response using the change in behavioural and neural positive bias as regressors of interest. Both analyses included Baseline HAM-D, Age, Gender, and use of Psychoactive Medication as control regressors.

### 4.11 Regression analysis including early clinical change

Six regression analyses were run to predict HAM-D Improvement, using early change in HAM-D (from Baseline to Week 2) and change in positive bias measures as regressors of interest. Baseline HAM-D, Age, Gender, Psychoactive Medication and Number of iTBS Sessions were included as control regressors. Each analysis included the change in one of the positive bias measures (change in rostral ACC response (Happy > Fear), precuneus and angular gyrus response (Happy > Fixation), PPI in precuneus and cuneus (Happy > Fear), reaction time for happy faces (fMRI task)). For the change in Positive Bias Misclassification, a logistic regression analysis was run predicting Clinical Response (since this measure was found to differ between Responders and Non-Responders).

### 4.12 Elastic net regression

To test whether the change in positive bias measures might improve prediction of HAM-D Improvement beyond early HAM-D change and demographic variables, two elastic net regression models were trained in Python (version 3.11.3) using *Scikit-Learn* (version 1.5.2). Elastic net is a regularised regression technique which combines L1 and L2 penalties to restrict the regression weights to avoid overfitting. The first model included early HAM-D change (Week 2), Baseline HAM-D, Age, Gender and Psychoactive Medication as predictor variables. The second model additionally included the change in all positive bias measures which were found to be related to clinical outcome (change in rostral ACC response (Happy > Fear), precuneus and angular gyrus response (Happy > Fixation), PPI in precuneus and cuneus (Happy > Fear), reaction time for happy faces (fMRI task), and in Positive Bias Misclassification (FERT)). The models were trained and evaluated in a nested cross-validation procedure using the *R^2^* score (coefficient of determination) as evaluation metric. In the outer cross-validation loop, the dataset was divided into 5 equal folds one of which was held back as test set. The remaining part of the dataset was used for hyperparameter tuning using conventional 5-fold cross-validation. The ‘L1 ratio’ parameter was optimised using grid search in the range of 0.1 to 1 in steps of 0.01. The model was then fitted to all 4 folds of the outer cross-validation loop using the best hyperparameter value, and model performance was evaluated on the test set. Since this test set was not involved in hyperparameter tuning, this technique provides a more accurate estimate of performance in independent datasets. Hyperparameter tuning was run five times so that each fold in the outer cross-validation loop served as test set once. The entire procedure was repeated 10 times, so that 50 estimates for model performance were obtained in total for each model.

### 4.1 Deviations from pre-registration

We decided to analyse the clinical outcome at Week 4 (instead of Week 6) to focus on the initial response to iTBS treatment rather than maintenance of the antidepressant effect. We included an additional positive bias measure in the analysis of the FERT, i.e. Positive Bias Misclassification. In contrast to the other positive bias measures, Positive Bias Misclassification captures a bias in response criterion, and has been shown to be sensitive to modulation by antidepressant treatment [22]. We added the ventral striatum as additional region of interest in the fMRI analysis since depression is associated with decreased response of the ventral striatum to rewards, and selective serotonin reuptake inhibitors have been shown to normalise activity in this region [66, 67]. In addition to our pre-registered analysis, we conducted a PPI analysis to assess changes in task-related connectivity which provides a more complete and interpretable picture of the effects of iTBS treatment. We added the regression analyses and machine learning models including early clinical change to test whether the change in positive bias measures predicted HAM-D Improvement beyond early clinical change.

## 5 Data availability statement

Analysis scripts are available at https://osf.io/fmhpt/. Due to the sensitive nature of our data, we are unable to share our data.

## 6 Disclosures

The authors declare that they have no conflicts of interests.

## Acknowledgements

We would like to thank Emile Radyte for her contributions to the analysis. This research was funded by a Newton International Advanced Fellowship (NAFR12/1010). JO’S is a Sir Henry Dale Fellow funded by the Royal Society and the Wellcome Trust (215451/Z/19/Z). The Wellcome Centre for Integrative Neuroimaging is supported by core funding from the Wellcome Trust (203139/Z/16/Z and 203139/A/16/Z). This project was supported by the NIHR Oxford Health Biomedical Research Centre (NIHR203316). The views expressed are those of the author(s) and not necessarily those of the NIHR or the Department of Health and Social Care. For the purpose of open access, the author has applied a CC BY public copyright license to any Author Accepted Manuscript version arising from this submission.

## Notes

### Competing Interest Statement

The authors have declared no competing interest.

### Clinical Trial

NCT04969549

### Clinical Protocols

https://osf.io/rhxm2

### Funding Statement

This research was funded by a Newton International Advanced Fellowship (NAFR12/1010). JOS is a Sir Henry Dale Fellow funded by theRoyal Society and the Wellcome Trust (215451/Z/19/Z). The Wellcome Centre for Integrative Neuroimaging is supported by core funding from the Wellcome Trust (203139/Z/16/Z and 203139/A/16/Z). This project was
supported by the NIHR Oxford Health Biomedical Research Centre (NIHR203316). The views expressed arethose of the author(s) and not necessarily those of the NIHR or the Department of Health and Social Care.

### Author Declarations

The study was approved by the National Ethics Committee in Brazil (CAEE: 52384921.6.0000.0068).

## References

1. World Health Organization (2017): Depression and other common mental disorders: global health estimates. World Health Organization.

2. Rush AJ, Trivedi MH, Wisniewski SR, Nierenberg AA, Stewart JW, Warden D, et al. (2006): Acute and longer-term outcomes in depressed outpatients requiring one or several treatment steps: a STAR* D report. American Journal of Psychiatry. 163:1905–1917.

3. McLachlan G (2018): Treatment resistant depression: what are the options? BMJ: British Medical Journal (Online). 363.

4. Berlim MT, van den Eynde F, Tovar-Perdomo S, Daskalakis ZJ (2014): Response, remission and drop-out rates following high-frequency repetitive transcranial magnetic stimulation (rTMS) for treating major depression: a systematic review and meta-analysis of randomized, double-blind and sham-controlled trials. Psychol Med. 44:225–239.

5. Blumberger DM, Vila-Rodriguez F, Thorpe KE, Feffer K, Noda Y, Giacobbe P, et al. (2018): Effectiveness of theta burst versus high-frequency repetitive transcranial magnetic stimulation in patients with depression (THREE-D): a randomised non-inferiority trial. The Lancet. 391:1683–1692.

6. Cole EJ, Phillips AL, Bentzley BS, Stimpson KH, Nejad R, Barmak F, et al. (2022): Stanford Neuromodulation Therapy (SNT): A Double-Blind Randomized Controlled Trial. Am J Psychiatry. 179:132–141.

7. Harmer CJ, Cowen PJ (2013): ’It’s the way that you look at it’--a cognitive neuropsychological account of SSRI action in depression. Philos Trans R Soc Lond B Biol Sci. 368:20120407.

8. Godlewska BR, Browning M, Norbury R, Cowen PJ, Harmer CJ (2016): Early changes in emotional processing as a marker of clinical response to SSRI treatment in depression. Transl Psychiatry. 6:e957.

9. Shiroma PR, Thuras P, Johns B, Lim KO (2014): Emotion recognition processing as early predictor of response to 8-week citalopram treatment in late-life depression. Int J Geriatr Psychiatry. 29:1132–1139.

10. Disner SG, Beevers CG, Haigh EA, Beck AT (2011): Neural mechanisms of the cognitive model of depression. Nat Rev Neurosci. 12:467–477.

11. Baeken C, Marinazzo D, Everaert H, Wu GR, Van Hove C, Audenaert K, et al. (2015): The Impact of Accelerated HF-rTMS on the Subgenual Anterior Cingulate Cortex in Refractory Unipolar Major Depression: Insights From 18FDG PET Brain Imaging. Brain Stimul. 8:808–815.

12. Eshel N, Keller CJ, Wu W, Jiang J, Mills-Finnerty C, Huemer J, et al. (2020): Global connectivity and local excitability changes underlie antidepressant effects of repetitive transcranial magnetic stimulation. Neuropsychopharmacology.

13. Fox MD, Buckner RL, White MP, Greicius MD, Pascual-Leone A (2012): Efficacy of transcranial magnetic stimulation targets for depression is related to intrinsic functional connectivity with the subgenual cingulate. Biol Psychiatry. 72:595–603.

14. Baeken C, Van Schuerbeek P, De Raedt R, De Mey J, Vanderhasselt MA, Bossuyt A, et al. (2010): The effect of one left-sided dorsolateral prefrontal cortical HF-rTMS session on emotional brain processes in women. Psychiatr Danub. 22 Suppl 1:S163.

15. Baeken C, De Raedt R, Van Schuerbeek P, Vanderhasselt MA, De Mey J, Bossuyt A, et al. (2010): Right prefrontal HF-rTMS attenuates right amygdala processing of negatively valenced emotional stimuli in healthy females. Behav Brain Res. 214:450–455.

16. Balconi M, Ferrari C (2013): Repeated transcranial magnetic stimulation on dorsolateral prefrontal cortex improves performance in emotional memory retrieval as a function of level of anxiety and stimulus valence. Psychiatry Clin Neurosci. 67:210–218.

17. Balconi M, Ferrari C (2012): rTMS stimulation on left DLPFC affects emotional cue retrieval as a function of anxiety level and gender. Depress Anxiety. 29:976–982.

18. Tozzi L, Zhang X, Pines A, Olmsted AM, Zhai ES, Anene ET, et al. (2024): Personalized brain circuit scores identify clinically distinct biotypes in depression and anxiety. Nat Med. 30:2076–2087.

19. Young AW, Rowland D, Calder AJ, Etcoff NL, Seth A, Perrett DI (1997): Facial expression megamix: Tests of dimensional and category accounts of emotion recognition. Cognition. 63:271–313.

20. Fu CH, Williams SC, Cleare AJ, Brammer MJ, Walsh ND, Kim J, et al. (2004): Attenuation of the neural response to sad faces in major depressionby antidepressant treatment: a prospective, event-related functional magnetic resonance imagingstudy. Archives of general psychiatry. 61:877–889.

21. Bhagwagar Z, Cowen PJ, Goodwin GM, Harmer CJ (2004): Normalization of enhanced fear recognition by acute SSRI treatment in subjects with a previous history of depression. American Journal of Psychiatry. 161:166–168.

22. Walsh AEL, Browning M, Drevets WC, Furey M, Harmer CJ (2018): Dissociable temporal effects of bupropion on behavioural measures of emotional and reward processing in depression. Philos Trans R Soc Lond B Biol Sci. 373.

23. Gur RC, Erwin RJ, Gur RE, Zwil AS, Heimberg C, Kraemer HC (1992): Facial emotion discrimination: II. Behavioral findings in depression. Psychiatry research. 42:241–251.

24. Harmer CJ, O’Sullivan U, Favaron E, Massey-Chase R, Ayres R, Reinecke A, et al. (2009): Effect of acute antidepressant administration on negative affective bias in depressed patients. American Journal of Psychiatry. 166:1178–1184.

25. Seeley WW, Menon V, Schatzberg AF, Keller J, Glover GH, Kenna H, et al. (2007): Dissociable intrinsic connectivity networks for salience processing and executive control. J Neurosci. 27:2349–2356.

26. Raichle ME, MacLeod AM, Snyder AZ, Powers WJ, Gusnard DA, Shulman GL (2001): A default mode of brain function. Proceedings of the national academy of sciences. 98:676–682.

27. Fox MD, Snyder AZ, Vincent JL, Corbetta M, Van Essen DC, Raichle ME (2005): The human brain is intrinsically organized into dynamic, anticorrelated functional networks. Proceedings of the National Academy of Sciences. 102:9673–9678.

28. Hamilton M (1960): A rating scale for depression. Journal of neurology, neurosurgery, and psychiatry. 23:56.

29. Park ASY, Thompson B (2024): Non-invasive brain stimulation and vision rehabilitation: a clinical perspective. Clinical and Experimental Optometry. 107:594–602.

30. Montgomery SA, Åsberg M (1979): A new depression scale designed to be sensitive to change. The British journal of psychiatry. 134:382–389.

31. Young RC, Biggs JT, Ziegler VE, Meyer DA (1978): A rating scale for mania: reliability, validity and sensitivity. The British journal of psychiatry. 133:429–435.

32. Watson D, Clark LA, Tellegen A (1988): Development and validation of brief measures of positive and negative affect: the PANAS scales. Journal of personality and social psychology. 54:1063.

33. Spielberger C (1983): Manual for the State-Trait Anxiety Inventory; Palo Alto, CA, Ed. Consulting Psychologists Press, Inc.: Columbia, MO, USA.

34. Grimm S, Boesiger P, Beck J, Schuepbach D, Bermpohl F, Walter M, et al. (2009): Altered negative BOLD responses in the default-mode network during emotion processing in depressed subjects. Neuropsychopharmacology. 34:932–943.

35. O’Reilly JX, Woolrich MW, Behrens TE, Smith SM, Johansen-Berg H (2012): Tools of the trade: psychophysiological interactions and functional connectivity. Soc Cogn Affect Neurosci. 7:604–609.

36. Finn ES (2021): Is it time to put rest to rest? Trends Cogn Sci. 25:1021–1032.

37. Pizzagalli DA (2011): Frontocingulate dysfunction in depression: toward biomarkers of treatment response. Neuropsychopharmacology. 36:183–206.

38. Boes AD, Uitermarkt BD, Albazron FM, Lan MJ, Liston C, Pascual-Leone A, et al. (2018): Rostral anterior cingulate cortex is a structural correlate of repetitive TMS treatment response in depression. Brain Stimul. 11:575–581.

39. Borserio BJ, Sharpley CF, Bitsika V, Sarmukadam K, Fourie PJ, Agnew LL (2021): Default mode network activity in depression subtypes. Rev Neurosci. 32:597–613.

40. Zhang B, Li S, Zhuo C, Li M, Safron A, Genz A, et al. (2017): Altered task-specific deactivation in the default mode network depends on valence in patients with major depressive disorder. J Affect Disord. 207:377–383.

41. Peng X, Wu X, Gong R, Yang R, Wang X, Zhu W, et al. (2021): Sub-regional anterior cingulate cortex functional connectivity revealed default network subsystem dysfunction in patients with major depressive disorder. Psychol Med. 51:1687–1695.

42. Zhou Z, Gao Y, Bao W, Liang K, Cao L, Tang M, et al. (2024): Distinctive intrinsic functional connectivity alterations of anterior cingulate cortex subdivisions in major depressive disorder: A systematic review and meta-analysis. Neurosci Biobehav Rev. 159:105583.

43. Margulies DS, Kelly AMC, Uddin LQ, Biswal BB, Castellanos FX, Milham MP (2007): Mapping the functional connectivity of anterior cingulate cortex. NeuroImage. 37:579–588.

44. Godlewska BR, Norbury R, Selvaraj S, Cowen PJ, Harmer CJ (2012): Short-term SSRI treatment normalises amygdala hyperactivity in depressed patients. Psychol Med. 42:2609–2617.

45. Ma Y (2015): Neuropsychological mechanism underlying antidepressant effect: a systematic meta-analysis. Mol Psychiatry. 20:311–319.

46. Murphy SE, Norbury R, O’Sullivan U, Cowen PJ, Harmer CJ (2009): Effect of a single dose of citalopram on amygdala response to emotional faces. Br J Psychiatry. 194:535–540.

47. Liston C, Chen AC, Zebley BD, Drysdale AT, Gordon R, Leuchter B, et al. (2014): Default mode network mechanisms of transcranial magnetic stimulation in depression. Biol Psychiatry. 76:517–526.

48. Philip NS, Barredo J, van ’t Wout-Frank M, Tyrka AR, Price LH, Carpenter LL (2018): Network Mechanisms of Clinical Response to Transcranial Magnetic Stimulation in Posttraumatic Stress Disorder and Major Depressive Disorder. Biol Psychiatry. 83:263–272.

49. Taylor SF, Ho SS, Abagis T, Angstadt M, Maixner DF, Welsh RC, et al. (2018): Changes in brain connectivity during a sham-controlled, transcranial magnetic stimulation trial for depression. J Affect Disord. 232:143–151.

50. Sheehan DV, Lecrubier Y, Sheehan KH, Amorim P, Janavs J, Weiller E, et al. (1998): The Mini-International Neuropsychiatric Interview (MINI): the development and validation of a structured diagnostic psychiatric interview for DSM-IV and ICD-10. Journal of clinical psychiatry. 59:22–33.

51. Altman DG, Royston P (2006): The cost of dichotomising continuous variables. Bmj. 332:1080.

52. Heeger D, Landy M (1997): Signal detection theory. *Dept Psych, Stanford Univ, Stanford, CA*, Teaching Handout.

53. R Core Team (2020): RA language and environment for statistical computing, R Foundation for Statistical. Computing.

54. Jenkinson M, Beckmann CF, Behrens TE, Woolrich MW, Smith SM (2012): FSL. Neuroimage. 62:782–790.

55. Smith SM (2002): Fast robust automated brain extraction. Hum Brain Mapp. 17:143–155.

56. Jenkinson M, Smith S (2001): A global optimisation method for robust affine registration of brain images. Medical image analysis. 5:143–156.

57. Jenkinson M, Bannister P, Brady M, Smith S (2002): Improved optimization for the robust and accurate linear registration and motion correction of brain images. Neuroimage. 17:825–841.

58. Gholipour A, Kehtarnavaz N, Gopinath K, Briggs R, Panahi I (2008): Average field map image template for Echo-Planar image analysis. 2008 30th Annual International Conference of the IEEE Engineering in Medicine and Biology Society: IEEE, pp 94-97.

59. Beckmann CF, Smith SM (2004): Probabilistic independent component analysis for functional magnetic resonance imaging. IEEE Trans Med Imaging. 23:137–152.

60. Griffanti L, Douaud G, Bijsterbosch J, Evangelisti S, Alfaro-Almagro F, Glasser MF, et al. (2017): Hand classification of fMRI ICA noise components. Neuroimage. 154:188–205.

61. Salimi-Khorshidi G, Douaud G, Beckmann CF, Glasser MF, Griffanti L, Smith SM (2014): Automatic denoising of functional MRI data: combining independent component analysis and hierarchical fusion of classifiers. Neuroimage. 90:449–468.

62. Griffanti L, Salimi-Khorshidi G, Beckmann CF, Auerbach EJ, Douaud G, Sexton CE, et al. (2014): ICA-based artefact removal and accelerated fMRI acquisition for improved resting state network imaging. Neuroimage. 95:232–247.

63. Andersson J, Jenkinson M, Smith S (2010): Non-linear registration, aka spatial normalisation. FMRIB technical report TR07JA2.

64. Winkler AM, Ridgway GR, Webster MA, Smith SM, Nichols TE (2014): Permutation inference for the general linear model. Neuroimage. 92:381–397.

65. Smith SM, Nichols TE (2009): Threshold-free cluster enhancement: addressing problems of smoothing, threshold dependence and localisation in cluster inference. Neuroimage. 44:83–98.

66. Pasquereau B, Drui G, Saga Y, Richard A, Millot M, Metereau E, et al. (2021): Selective serotonin reuptake inhibitor treatment retunes emotional valence in primate ventral striatum. Neuropsychopharmacology. 46:2073–2082.

67. Stoy M, Schlagenhauf F, Sterzer P, Bermpohl F, Hagele C, Suchotzki K, et al. (2012): Hyporeactivity of ventral striatum towards incentive stimuli in unmedicated depressed patients normalizes after treatment with escitalopram. J Psychopharmacol. 26:677–688.

