## Supplementary Material for "Transcranial Magnetic Stimulation Depression Treatment Induces Positive Bias in Task-Related Brain Function: Results From the BRAEN-MAP Trial"

#### Table of Contents

*1 Facial Expression Recognition Task (FERT) ... 2*

*2 Task fMRI ... 3*

**2.1 FMRI acquisition ... 3**

**2.2 TMS induces positive bias in neural response to emotional faces ... 4**

**2.3 TMS induces positive bias in task-related connectivity ... 7**

**2.4 Task performance ... 8**

2.4.1 Accuracy ... 8

2.4.2 Reaction Times ... 9

**2.5 Motion ... 11**

*3 Combined behavioural and neural analysis ... 13*

**3.1 Elastic net regression ... 13**

*4 Supplementary tables ... 15*

### 1 Facial Expression Recognition Task (FERT)

As described in the main manuscript, Responders compared to Non-Responders showed an increase in Positive Bias Misclassification from Baseline to Week 2 (Figure S1A-B). Change in any of the four positive bias measures did not predict HAM-D Improvement as continuous variable (Positive Bias RT: $\beta = 0.76$, $t(39) = 0.11$, $p = .91$; Positive Bias UBH: $\beta = -2.45$, $t(39) = -0.66$, $p = .51$; Positive Bias Efficiency: $\beta = -4.22$, $t(39) = -1.43$, $p = .16$; Positive Bias Misclassification: $\beta = 2.36$, $t(39) = 1.67$, $p = .103$)(Figure S2). However, on a descriptive level, there was a positive relationship between change in Positive Bias Misclassification and HAM-D improvement, which is in line with the increase in positive bias in Responders compared to Non-Responders reported in the main text.

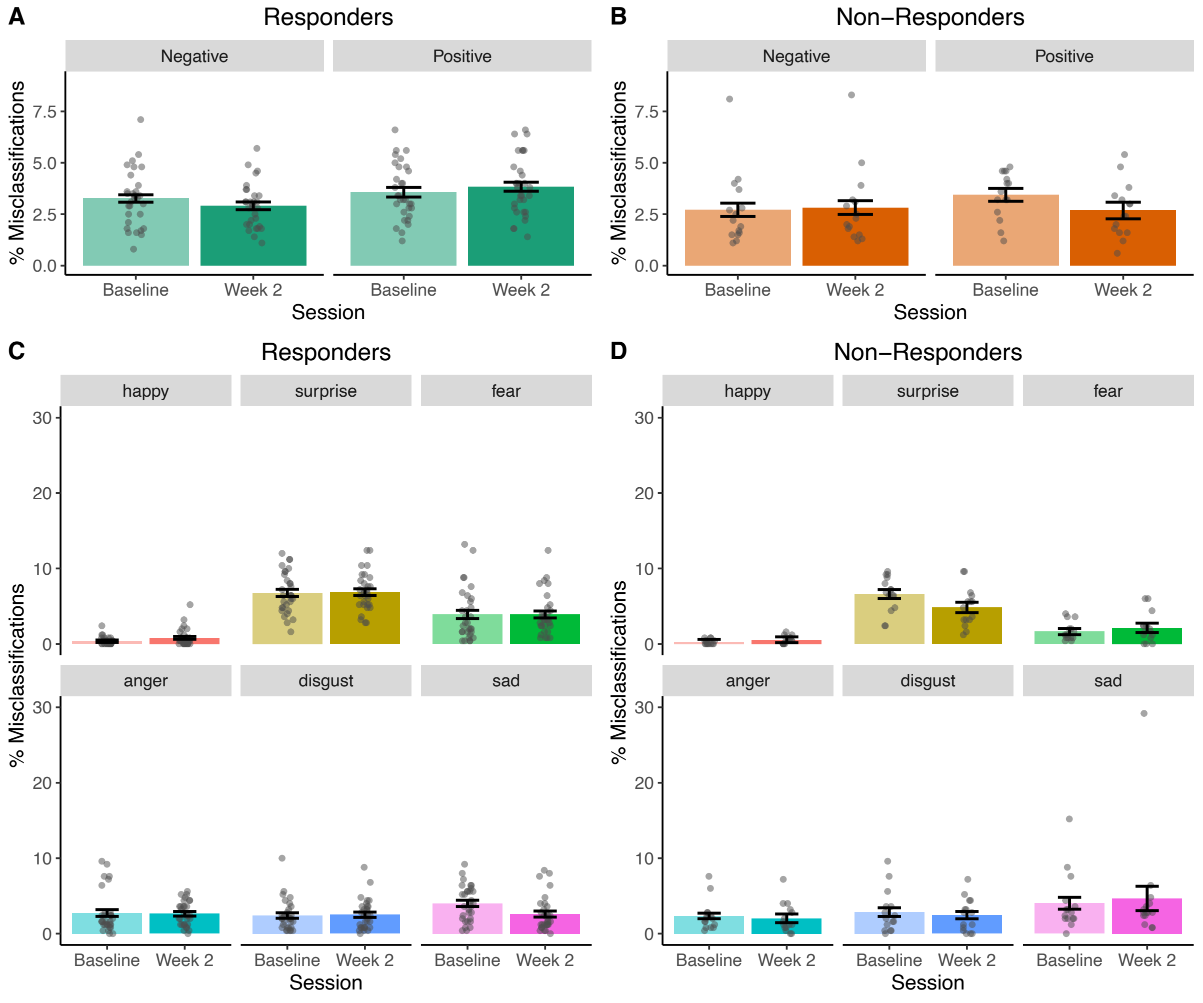


*Figure S1*. (A) and (B) show how misclassifications for positive and negative emotions changed in Responders (A) and Non-Responders (B) from Baseline to Week 2. (C) and (D) show the change in misclassifications for each emotion in Responders (C) and Non-Responders (D). The results suggest that the change in Positive Bias Misclassification was based on a change in the overall pattern in the data rather than a specific emotion. On a descriptive level, the effect was driven by a relative decrease in misclassifications as sadness in Responders, and a relative decrease in misclassifications as surprise in Non-Responders. Error bars represent SEM.

A follow-up ANOVA was run to test whether the increase in Positive Bias Misclassification was driven by a change in a particular emotion. There was a significant interaction effect between Clinical Response, Time and Emotion ($F$(5,220) = 2.4, $p$ = .045). Post-hoc ANOVAs for individual emotions did not show any significant interactions between Clinical Response and Time (all $p_{corr}$ > .25), suggesting that the change in Positive Bias Misclassification was based on a change in the overall pattern in the data rather than a specific emotion. On a descriptive level, the effect was driven by a relative decrease in misclassifications as sadness in Responders (F(1,44) = 4.3, $p$ = .042 (uncorrected)), and a relative decrease in misclassifications as surprise in Non-Responders (F(1,44) = 3.4, $p$ = .069 (uncorrected))(Figure S1C-D).

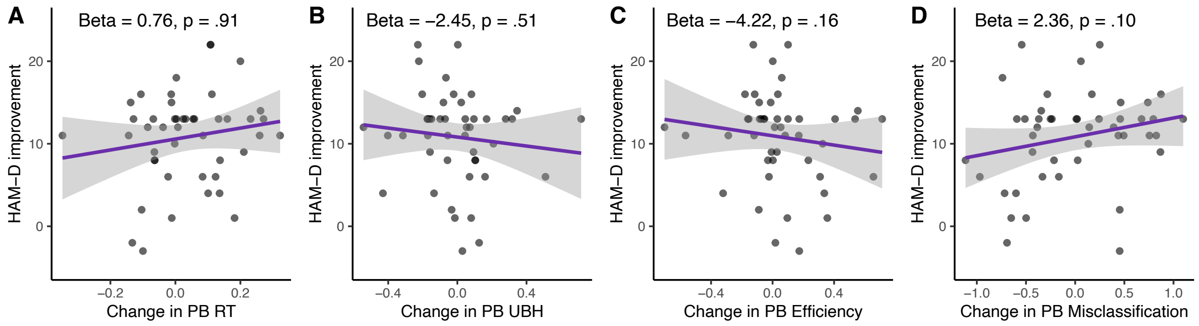


*Figure S2.* Relationship between early changes in positive bias in the Facial Expression Recognition Task (FERT) and HAM-D Improvement at the end of treatment. Early changes in Positive Bias RT (A), Positive Bias Unbiased Hit Rate (B), Positive Bias Efficiency (C) and Positive Bias Misclassification (D) did not predict HAM-D Improvement at the end of treatment. ‘ indicates a trend-level effect (.05 < $p$ < .10). ** indicates .001 < $p$ < .01. PB = Positive Bias, RT = Reaction Time, UBH = Unbiased Hit Rate. Error bars represent SEM.

### 2 Task fMRI

#### 2.1 FMRI acquisition

The scanner protocol was adapted from a UK Biobank protocol. Participants were scanned in a 3T Philips Achieva MRI scanner using echo-planar imaging. The voxel size was 3x3x3mm. TR and TE were 2.49s and 30ms, respectively, with a flip angle of 90°. The field of view was 240x240x120mm. The scan lasted 8.5mins with an acquisition of 204 volumes. Structural T1-weighted images were acquired with a resolution of 0.46x0.46x2mm, TR of 7ms, TE of 3.2ms and inversion time of 900ms. The field maps were acquired with a voxel size of 3x3x3mm, field of view of 240x240x120mm, flip angle of 50°, TR of 239ms, and TE of 2.3ms.

#### 2.2 TMS induces positive bias in neural response to emotional faces

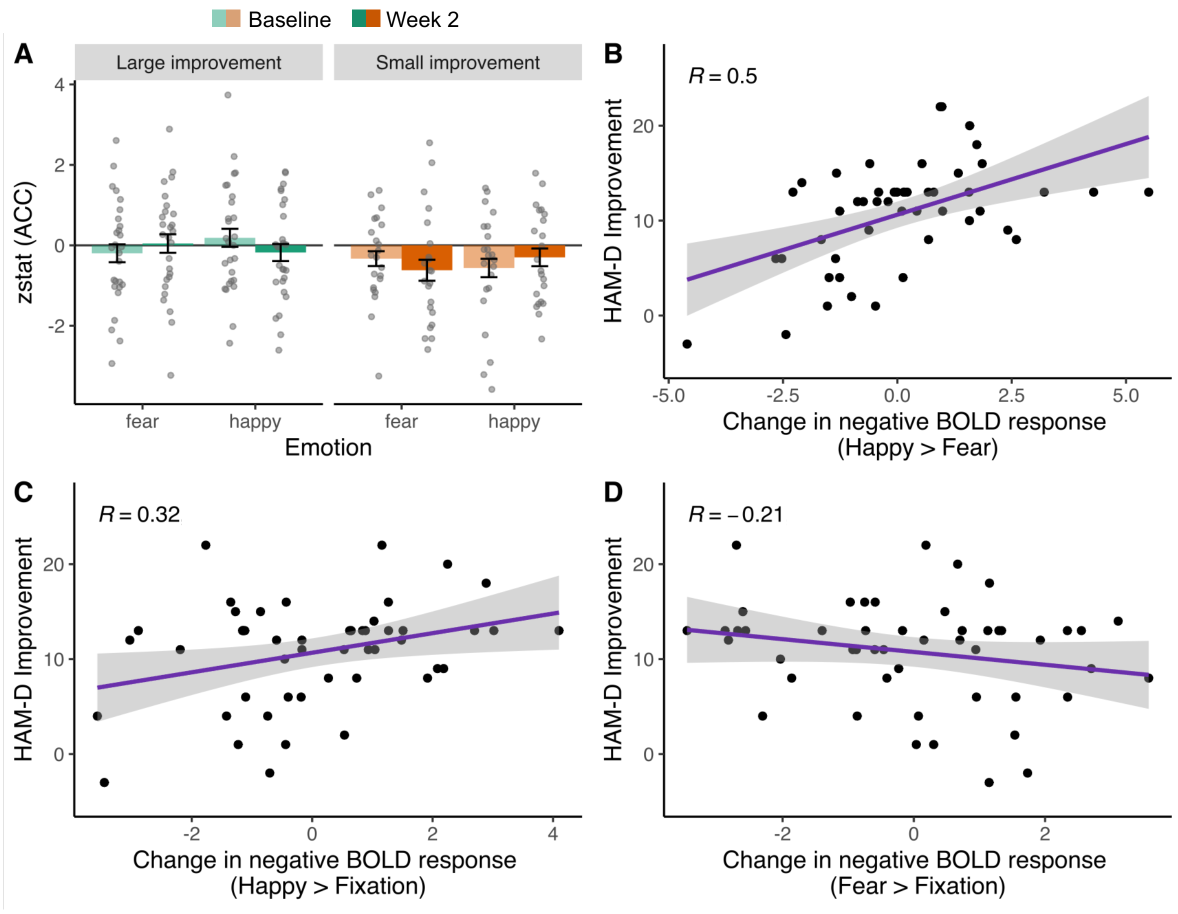


*Figure S3.* Visualisation of the correlation between HAM-D Improvement and increase in negative BOLD response for Happy > Fearful faces in the rostral ACC. (A) shows a median split for HAM-D Improvement which was performed solely for visualisation purposes. (B) shows the correlation between HAM-D Improvement and increase in negative BOLD response for Happy > Fearful faces. (C) and (D) show the same correlation separately for the constituent contrasts Happy > Fixation and Fear > Fixation. These visualisations suggest that the effect was primarily driven by changes in the response to happy faces. Error bars represent SEM. Please note that a positive value for 'change in negative BOLD response' indicates an increase in negative BOLD response (i.e. more negative BOLD signal).

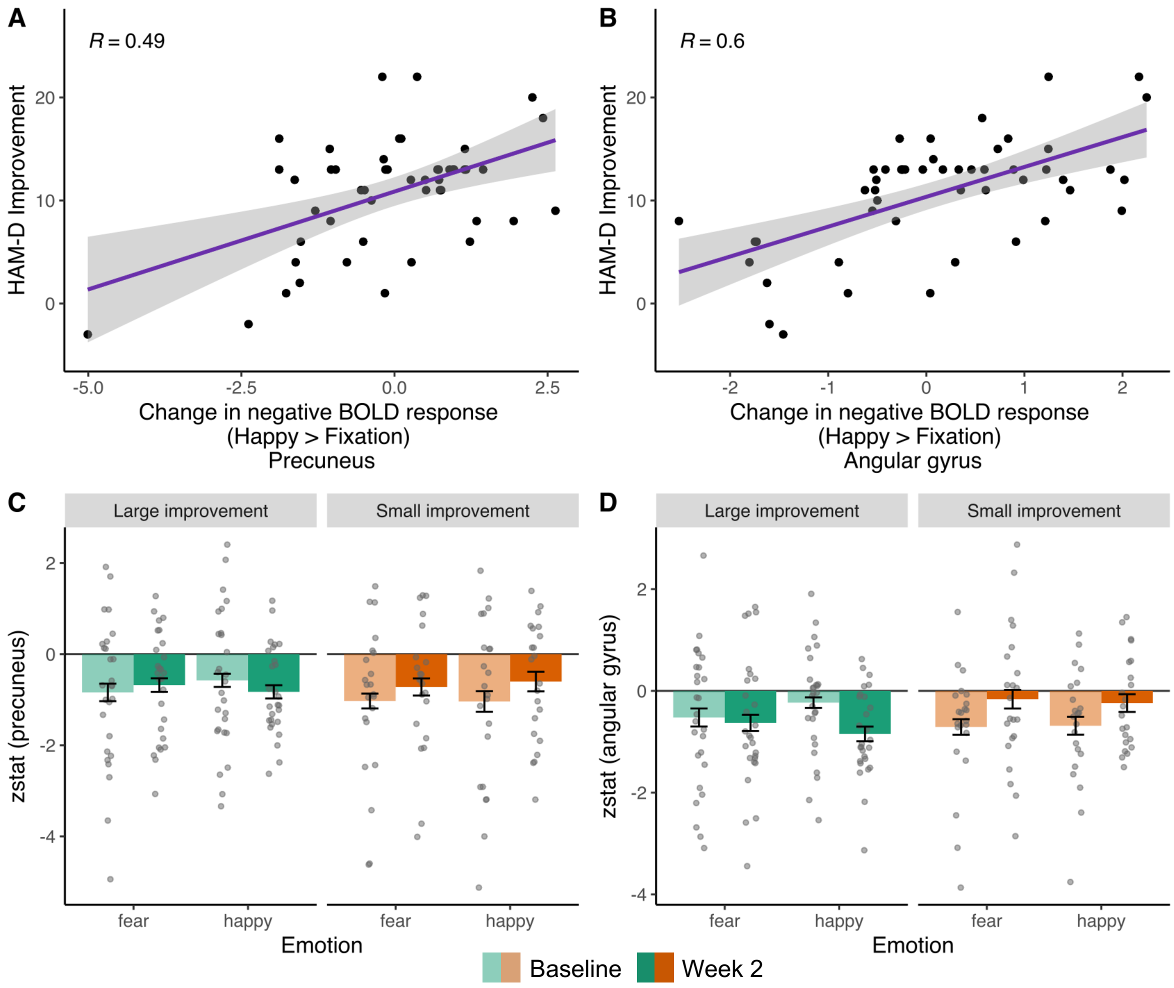


*Figure S4.* Visualisation of the relationship between HAM-D Improvement and increase in negative BOLD response to happy faces (Happy > Fixation). (A) and (B) show the correlation between HAM-D Improvement and change in negative BOLD signal in the Happy > Fixation contrast in the precuneus and left angular gyrus. (C) and (D) show the result of a median split based on HAM-D improvement which was performed solely for visualisation purposes. Individuals with larger HAM-D improvement showed a relative increase in negative BOLD responses to happy faces. Error bars represent SEM. Please note that a positive value for 'change in negative BOLD response' indicates an increase in negative BOLD response (i.e. more negative BOLD signal).

The analyses reported in the main text included HAM-D Improvement as continuous regressor. When repeating the analysis above including Clinical Response (instead of HAM-D Improvement) as categorical regressor, no significant clusters emerged in the Happy > Fearful contrast on the whole-brain level. ROI analysis revealed a cluster in the rostral ACC which overlapped with the one found in the analysis on HAM-D Improvement, but it did not survive multiple comparison correction (threshold 0.99). The whole-brain analysis for Happy > Fixation revealed a significant cluster for the Non-Responders > Responders contrast in the left angular gyrus (159 voxels, $t_{max} = 5.15$, $p = .026$, x = -58, y = -66, z = 22), i.e. Responders showed a larger increase is negative BOLD response to happy faces than Non-Responders (Figure S4). This cluster overlaps with the one observed for HAM-D Improvement. No significant clusters were found in the Fearful > Fixation contrast.

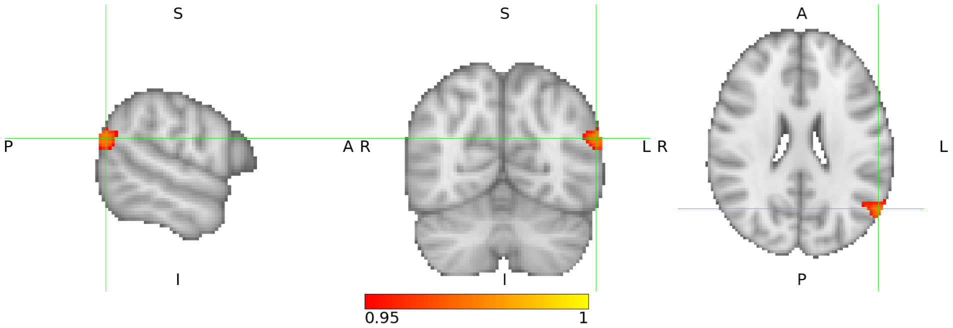


*Figure S5.* Responders vs. Non-Responders showed a larger increase in negative BOLD response to happy faces in the angular gyrus. This is in line with the result from the analysis on HAM-D Improvement reported in the main manuscript.

#### 2.3 TMS induces positive bias in task-related connectivity

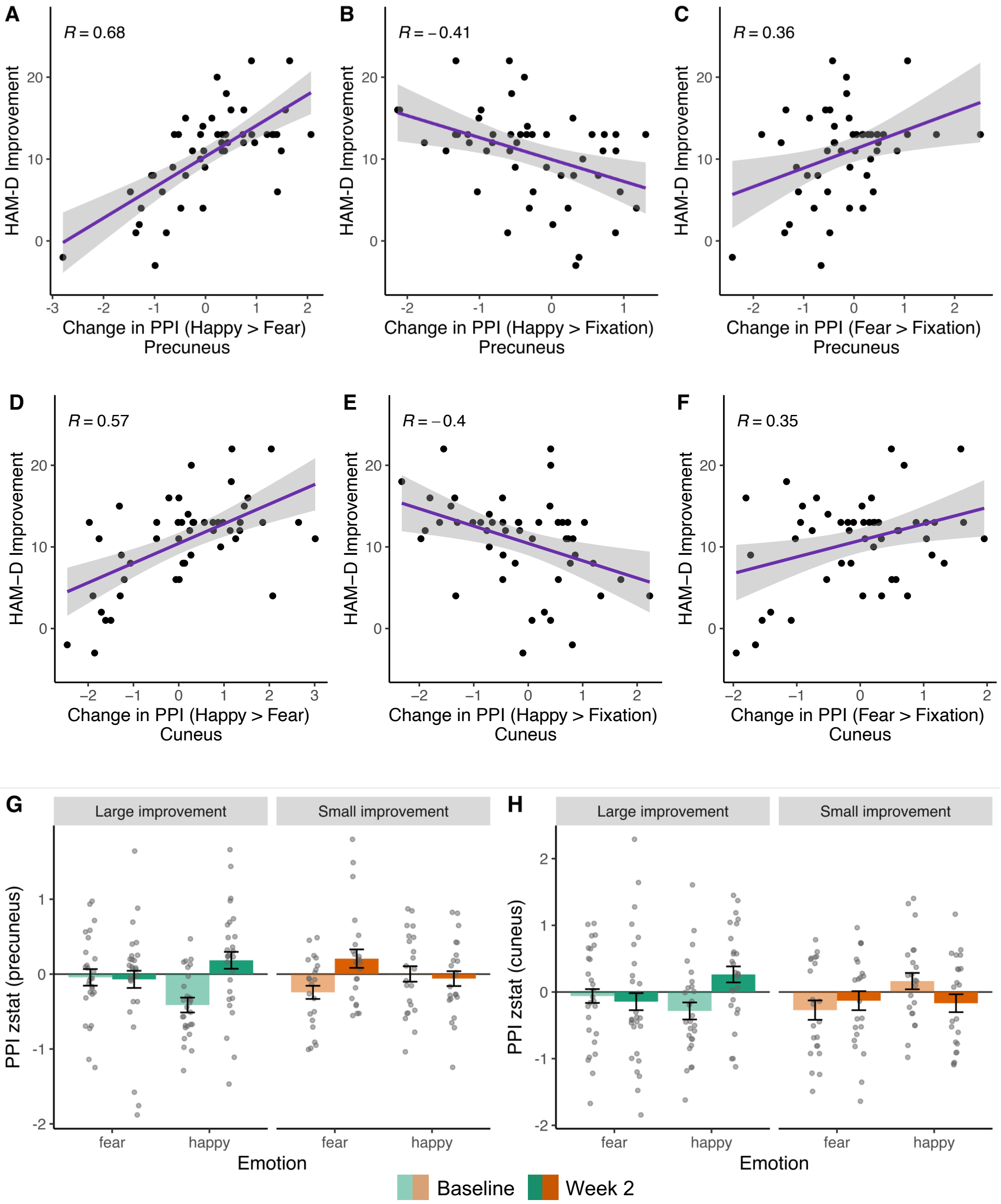


*Figure S6*. Visualisation of the psycho-physiological interaction (PPI) analysis including rostral ACC as seed region. Significant clusters in the Happy > Fearful contrast represent brain regions that showed a stronger connectivity with rostral ACC during the presentation of happy compared to fearful faces. The analysis revealed a positive correlation between HAM-D Improvement and increase in PPI in the Happy > Fearful contrast in several regions including precuneus (A-C, G) and cuneus (D-F, H). The scatter plots suggest that the observed correlations between increase in the Happy > Fearful contrast and HAM-D Improvement in precuneus and posterior cingulate were equally driven by changes in connectivity in the Fearful > Baseline and Happy > Baseline contrasts. The bar graphs (G, H) show PPI values (connectivity for Fear > Fixation and Happy > Fixation) for individuals with large or small HAM-D Improvement based on a median split. Error bars represent SEM.

#### 2.4 Task performance

This analysis included data of 42 participants (due to technical problems, responses could not be recorded for three participants; four participants were excluded from this analysis since they performed below chance performance (most likely due to confusion of the buttons)).

##### 2.4.1 Accuracy

Accuracy was defined as the percentage of correct responses in the gender discrimination task. To test whether accuracy changed over time as a function of Clinical Response, a mixed ANOVA was conducted including Session (Baseline vs. Week 2), Emotion (happy vs. fearful) and Clinical Response (Responders vs. Non-Responders). There was a significant main effect of Emotion on accuracy ($F(1,40) = 14.1$, $p < .001$). As shown in Figure S3, participants had a higher percentage of correct responses for fearful than for happy faces. No other main or interaction effect reached significance (all $p > .16$). This suggests that Responders and Non-Responders did not significantly differ in accuracy (or change in accuracy). In addition, two regression analyses were run to test whether the change in accuracy for happy faces or fearful faces predicted HAM-D Improvement as continuous variable (controlling for Baseline HAM-D, Age, Gender, Psychoactive Medication and Number of TMS Sessions). The change in task performance for happy or fearful faces did not predict HAM-D Improvement (Happy: $\beta = -0.17$, $t(35) = -1.2$, $p = .24$; Fearful: $\beta = -0.02$, $t(35) = -0.13$, $p = .90$).

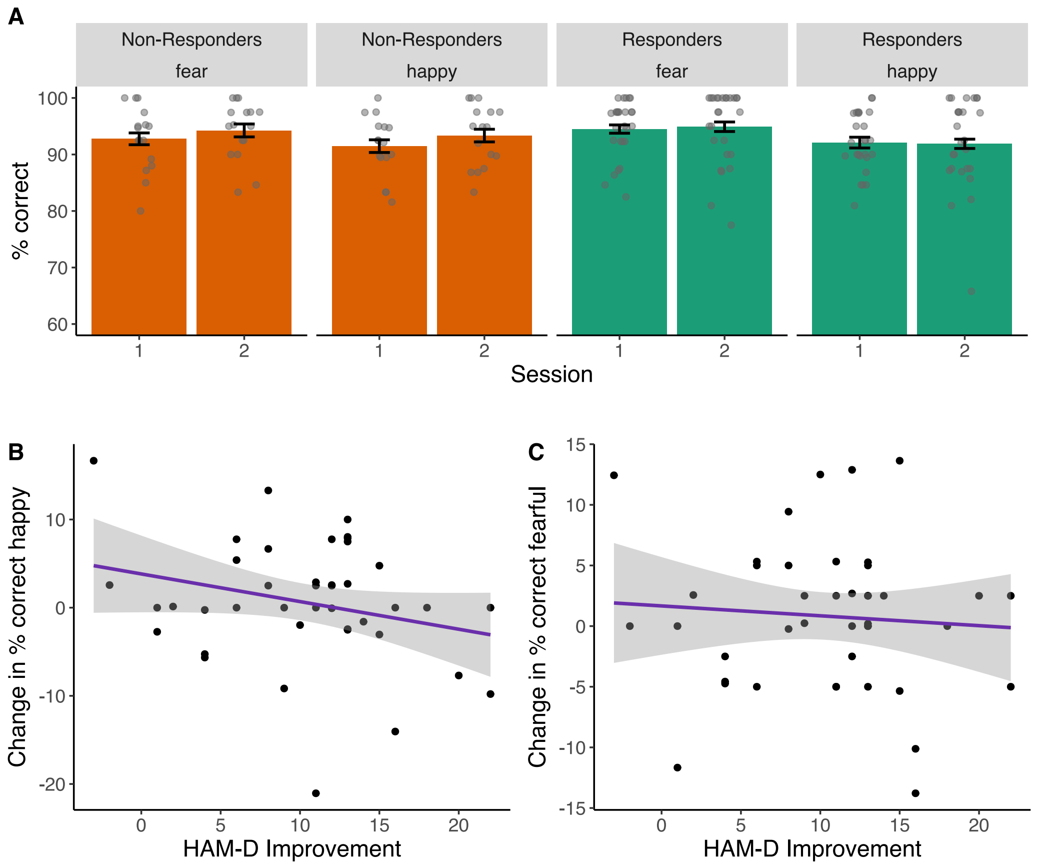


*Figure S7.* Accuracy in the fMRI gender discrimination task. (A) Clinical Response had no significant effect on task performance. Averaged across the groups, performance was higher for fearful than for happy faces. (B) HAM-D Improvement did not correlate with the change in performance for happy or fearful faces. Error bars represent SEM.

##### 2.4.2 Reaction Times

For each participant, the median reaction time (RT) was calculated separately for happy faces and fearful faces. This only included trials in which the gender was correctly identified. An ANOVA was conducted including RT as dependent variable, and Session (Baseline vs. Week 2), Emotion (happy vs. fearful) and Clinical Response (Responders vs. Non-Responders) as independent variables. There was an interaction effect between Clinical Response and Session ($F(1,40) = 5.31$, $p = .026$), and a three-way interaction effect between Clinical Response, Session and Emotion ($F(1,40) = 4.14$, $p = .0.048$). A follow-up ANOVA in Responders revealed a trend-level interaction between Session and Emotion ($F(1,25) = 4.07$, $p = .054$), indicating that reaction times for happy and fearful faces changed differently over time. However, the change was neither significant for happy ($F(1,25) = 2.65$, $p = 0.11$) nor for fearful faces ($F(1,25) =0.02$, $p = .88$). In Non-Responders, there was a trend-level main effect of Session ($F(1,15) = 3.84$, $p = .068$), indicating faster RT in the second session, but no other significant effects ($p > .32$).

In addition, two regression analyses were run to test whether the change in RT for happy faces or fearful faces predicted HAM-D Improvement as continuous variable (controlling for Baseline HAM-D, Age, Gender, Psychoactive Medication and Number of TMS Sessions). The change in RT for happy faces was a significant predictor for HAM-D Improvement (Happy: $\beta = 0.015$, $t(35) = 2.03$, $p = .0491$), such that a larger increase in RT from Baseline to Session 2 predicted larger HAM-D Improvement. The change in RT for fearful faces did not predict HAM-D Improvement (Fearful: $\beta = 0.011$, $t(35) = 1.33$, $p = .19$).

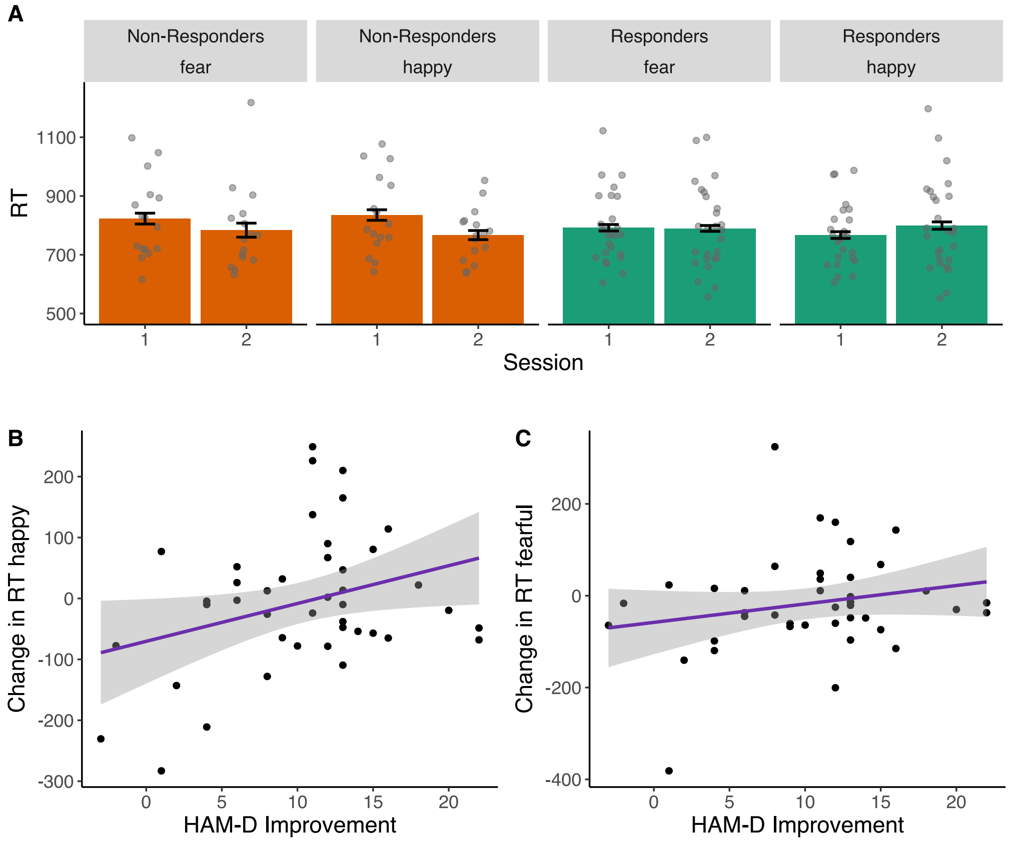


*Figure S8.* Reaction times in the fMRI gender discrimination task. (A) Non-Responders showed a trend-level effect towards shorter reaction times in Session 2 compared to the Baseline Session. (B) An increase in reaction times for happy faces predicted larger HAM-D Improvement. (C) No relationship between change in reaction time for fearful faces and HAM-D Improvement was found.

An increase in RT for happy faces could be interpreted as attentional capture, i.e. the happy facial expressions might distract from the task. This would be in line with an increase in positive bias. To test whether the increase in RT for happy faces was related to the increase in positive bias in brain activity and connectivity observed in the fMRI analysis, we calculated correlations between the change in RT for happy faces with the change in brain activity and connectivity reported in the main text. The increase in RT for happy faces correlated positively with the increase in response to happy faces in the precuneus ($r = 0.4$, $t(40) = 2.7$, $p = .0083$), and with the increase in connectivity (PPI) for happy vs. fearful faces in the precuneus ($r = 0.41$, $t(40) = 2.8$, $p = .0073$) and cuneus ($r = 0.5$, $t(40) = 3.6$, $p = .0007$) (Figure SX). The change in RT was not related to the increase in activity for happy vs. fearful faces in the rostral ACC ($r = 0.17$, $t(40) = 1.1$, $p = .28$) or the increase in response to happy faces in the angular gyrus ($r = 0.19$, $t(40) = 1.2$, $p = .24$).

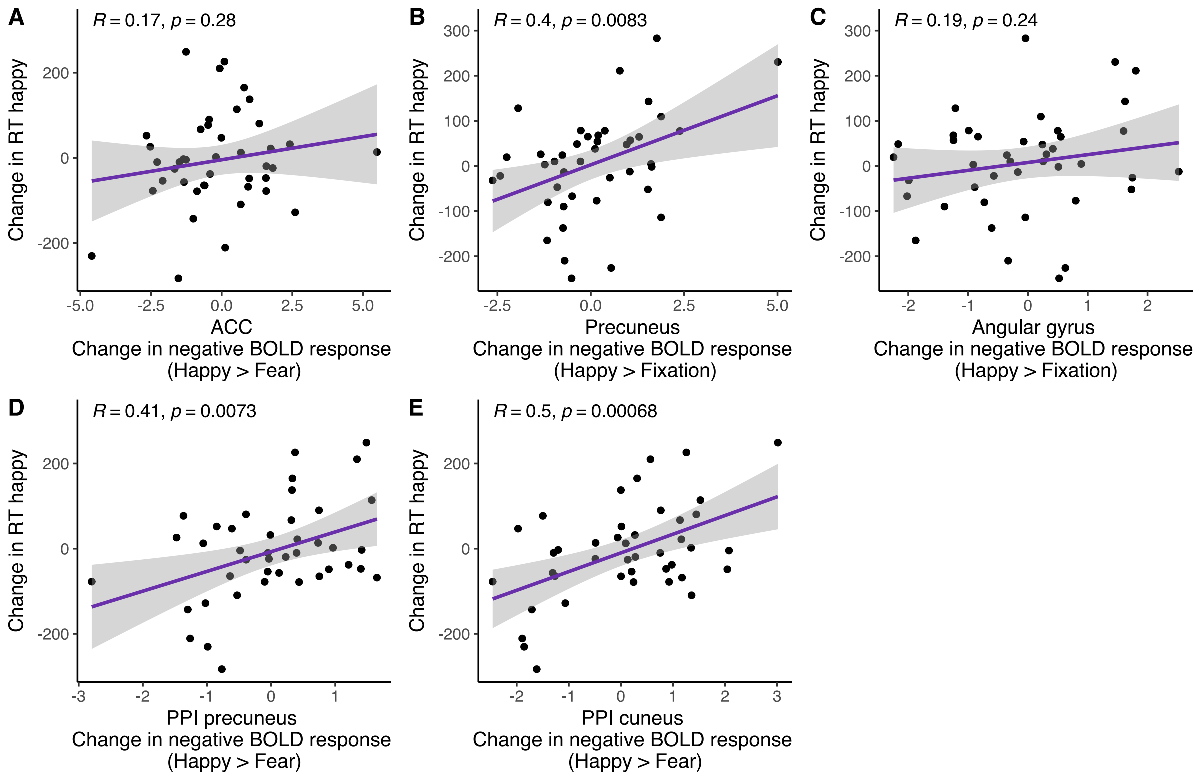


*Figure S9.* Correlations between the increase in RT for happy faces and the fMRI measures reported in the main text. The increase in RT was positively correlated with the increased response to happy faces in the precuneus (B), and increased connectivity for happy vs. fearful faces in the precuneus (D) and cuneus (E). The increase in activity to happy vs. fearful faces in the rostral ACC (A) and increase in response to happy faces in the angular gyrus (C) were not related to the increase in RT for happy faces. Please note that a positive value for 'change in negative BOLD response' indicates an increase in negative BOLD response (i.e. more negative BOLD signal).

#### 2.5 Motion

Absolute and relative motion estimates were extracted from FSL for each participant and each session. To test whether motion might be a confounding factor in our fMRI analysis, we tested whether motion changed from Baseline to Session 2, and whether motion (and the change in motion) was related to the clinical outcome. We conducted two ANOVAs with relative motion or absolute motion as dependent variable, and Session (Baseline vs. Week 2) and Clinical Response as independent variables. There were no significant effects for absolute or relative motion, indicating that motion did not change over time, and did not differ between Responders and Non-Responders (Absolute motion: Clinical Response: $F(1,47) = 1.73$, $p = .19$; Session: $F(1,47) = 0.43$, $p = .51$; Clinical Response x Session: $F(1,47) = 0.57$, $p = .45$; Relative motion: Clinical Response: $F(1,47) = 1.07$, $p = .30$; Session: $F(1,47) = 1.00$, $p = .32$; Clinical Response x Session: $F(1,47) = 0.20$, $p = .65$).

Moreover, we ran two regression analyses to test whether change in absolute or relative motion was related to change in HAM-D score. HAM-D Improvement was included as dependent variable, and change in absolute motion or change in relative motion were included as regressors of interest. Baseline HAM-D, Age, Gender, Psychoactive Medication and Number of TMS Sessions were included as control regressors. Neither the change in absolute motion nor the change in relative motion were significantly related to HAM-D Improvement

(Change in absolute motion: $\beta$ = -0.11, $t(42)$ = -0.04, $p$ = .97; Change in relative motion: $\beta$ = -9.95, $t(42)$ = -0.51, $p$ = .60). Taken together, there was no evidence for relationship between amount of motion and clinical outcome, which suggests that the results from our task fMRI analysis are unlikely to be confounded by motion.

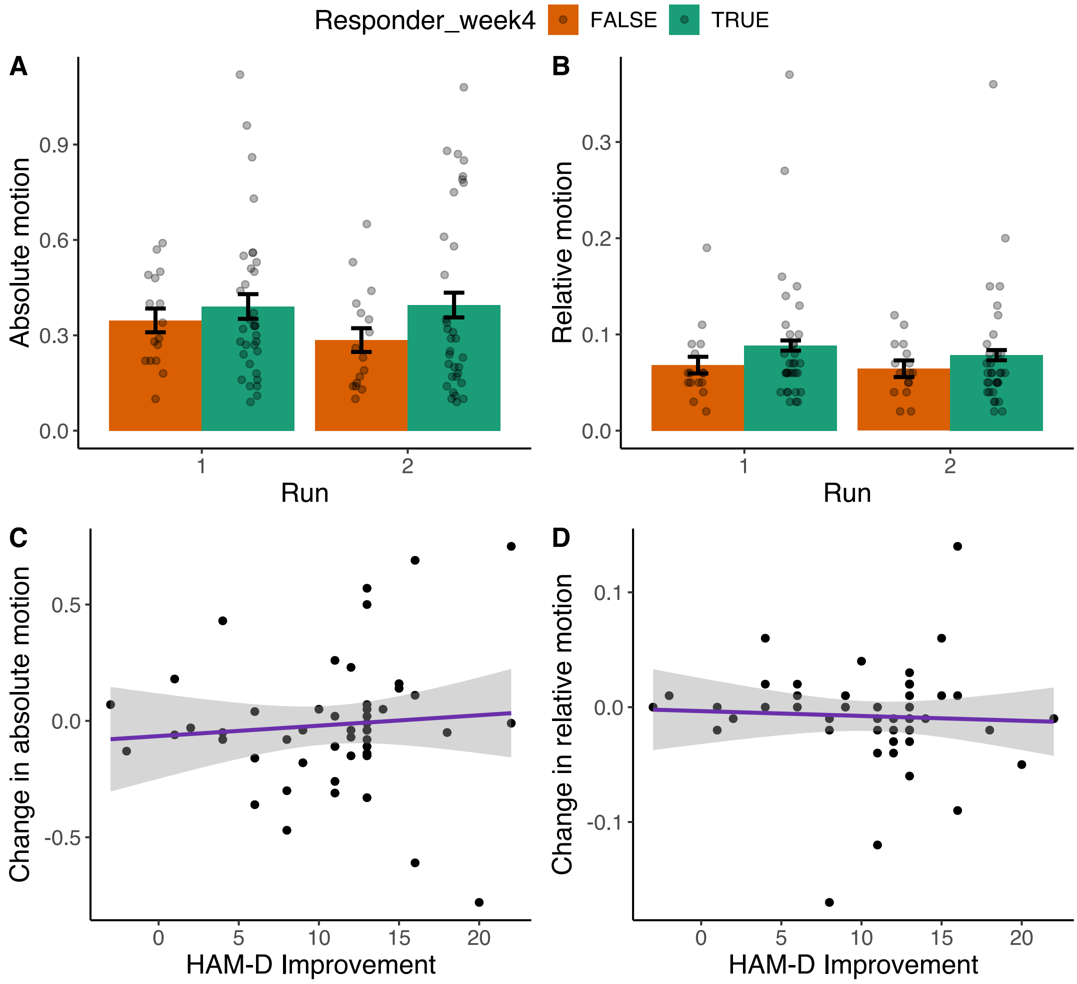


*Figure S10.* Absolute and relative motion estimates during the fMRI scans. Responders and Non-Responders did not differ in absolute (A) or relative motion (B). Motion did not differ between sessions (run 1 and 2). The change in absolute (C) or relative motion (D) between Baseline and Week 2 was not related to HAM-D Improvement. This indicates that our fMRI results are unlikely to be confounded by motion.

### 3 Combined behavioural and neural analysis

#### 3.1 Elastic net regression

To test whether the change in positive bias measures might improve prediction of HAM-D Improvement beyond early HAM-D change and demographic variables, two elastic net regression models were trained in Python (version 3.11.3) using *Scikit-Learn* (version 1.5.2). Elastic net is a regularised regression technique which combines L1 and L2 penalties to restrict the regression weights to avoid overfitting. The L2 penalty term shrinks the weights towards one another, whereas the L1 penalty term reduces weights of features with low importance to zero. The 'L1 ratio' hyperparameter determines the strength of L1 vs. L2 penalty.

The Baseline Model included early HAM-D change (Week 2), Baseline HAM-D, Age, Gender and Psychoactive Medication as predictor variables. The Bias Change Model additionally included the change in all positive bias measures which were found to be related to clinical outcome (change in rostral ACC response (Happy > Fear), precuneus and angular gyrus response (Happy > Fixation), PPI in precuneus and cuneus (Happy > Fear), reaction time for happy faces (fMRI task), and in Positive Bias Misclassification (FERT)).

The models were trained and evaluated in a nested cross-validation procedure. In the outer cross-validation loop, the dataset was divided into 5 equal folds one of which was held back as test set. The remaining part of the dataset was used for hyperparameter tuning using conventional 5-fold cross-validation, i.e. the remaining data were again split into 5 equal folds, and the model was fitted to four training folds using each possible hyperparameter value, and evaluated on the fifth fold serving as validation set. The 'L1 ratio' hyperparameter was optimised using grid search in the range of 0.1 to 1 in steps of 0.01. This was repeated 5 times so that each fold was used as validation set once. The model was then fitted to all four training folds in the outer cross-validation loop using the hyperparameter value that yielded the best average validation performance ($R^2$ score, coefficient of determination) in the inner cross-validation. The performance of this model was then evaluated on the test set, which provides a more accurate estimate of performance in independent datasets since the test set was not used for hyperparameter tuning. The inner cross-validation loop was run five times so that each fold in the outer cross-validation loop served as test set once. The entire procedure was repeated 10 times, so that 50 estimates for model performance were obtained in total for each model (Figure S11). The mean and standard deviation of these 50 estimates were calculated as performance metrics.

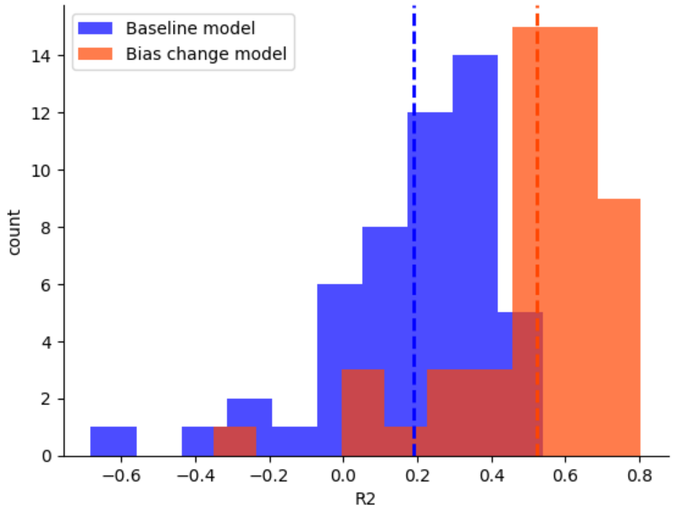


*Figure S11. $R^2$* scores from the nested cross-validation procedure for the Baseline model and the Bias change model (which included the change in behavioural and neural markers of positive bias in addition to the predictors of the Baseline model). The vertical lines represent the mean of the two distributions. The Bias Change Model clearly outperformed the Baseline Model.

Figure S12 displays the regression coefficients for each model after refitting the models using the best hyperparameter values. Due to the L1 penalty, elastic net regression tends to set coefficients of features with low importance to zero. Since the coefficients for the two behavioural positive bias measures were set to zero, this confirms the results of our analyses reported in the main text suggesting that the change in these measures does not predict HAM-D Improvement beyond early HAM-D change (Week2). All neural measures of positive bias, however, were assigned non-zero weights, confirming their predictive potential.

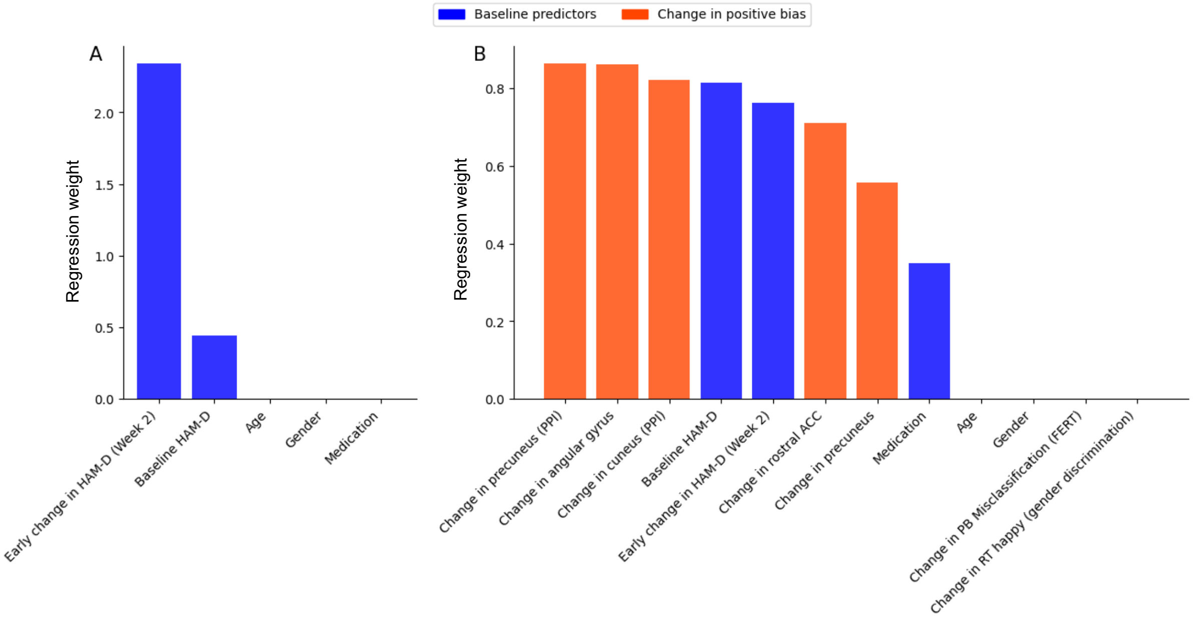


*Figure S12.* Regression weights for the two elastic net models. (A) shows the regression weights for the Baseline Model containing demographic data and early change in HAM-D score. (B) shows the weights for the Bias Change Model that also included the change in neural and behavioural markers of positive bias. Please not that some features were assigned a weight of zero (L1 penalty).

### 4 Supplementary tables

**Table S1. Psychoactive medication which participants were taking during study participation.**

| Type | Drug | Number of participants |
|---|---|---|
| SSRI | Escitalopram | 2 |
| | Fluoxetine | 2 |
| | Sertraline | 3 |
| SNRI | Desvenlafaxine | 7 |
| | Duloxetine | 2 |
| | Venlafaxine | 3 |
| Other antidepressants | Bupropion | 3 |
| | Tranylcypromine | 1 |
| | Trazodone | 2 |
| | Nortriptyline | 1 |
| | Amitriptyline | 2 |
| | Mirtazapine | 1 |
| Benzodiazepines | Alprazolam | 4 |
| | Clonazepam | 3 |
| Other anxiolytics | Pregabalin | 2 |
| Mood stabilisers | Lithium | 2 |
| | Lamotrigine | 1 |
| Antipsychotics | Brexpiprazole | 1 |
| | Olanzapine | 1 |
| | Chlorpromazine | 1 |
| | Aripiprazole | 1 |
| Other psychoactive medication | Lisdexamfetamine | 1 |
| | Methylfenidate | 2 |
| | Zolpidem | 2 |
| | Topiramate | 1 |

**Table S2. Percentage HAM-D change from Baseline for Responders and Non-Responders.**

| | Responders | Non-Responders |
|---|---|---|
| %-HAM-D change Week 1, mean (SD) | -33.8 (0.24) | -19.2 (0.24) |
| %-HAM-D change Week 2, mean (SD) | -55.7 (0.21) | -24.3 (0.14) |
| %-HAM-D change Week 3, mean (SD) | -61.4 (0.16) | -20.6 (0.27) |
| %-HAM-D change Week 4, mean (SD) | -75.4 (0.12) | -25.3 (0.22) |
| %-HAM-D change Week 6, mean (SD) | -65.1 (0.25) | -37.1 (0.25) |

**Table S3. Change in positive bias measures (Facial Expression Recognition Task) between Baseline and Week 2 for Responders and Non-Responders.**

| | Responders | | Non-Responders | |
|---|---|---|---|---|
| | Baseline | Week 2 | Baseline | Week 2 |
| Positive Bias Reaction Time, mean (SD) | 0.21 (0.12) | 0.26 (0.12) | 0.20 (0.14) | 0.23 (0.12) |
| Positive Bias Unbiased Hit Rate, mean (SD) | 0.56 (0.27) | 0.53 (0.29) | 0.43 (0.23) | 0.47 (0.21) |
| Positive Bias Efficiency, mean (SD) | 0.71 (0.30) | 0.72 (0.31) | 0.50 (0.16) | 0.64 (0.25) |
| Positive Bias Miscalssification, mean (SD) | 0.11 (0.56) | 0.27 (0.52) | 0.32 (0.59) | -0.03 (0.52) |

**Table S4. Peak activation for the task-negative network. X, Y, and Z coordinates are in MNI space (mm).**

| Cluster Index | Voxels | Brain regions | X | Y | Z | t-value | p-value |
|---|---|---|---|---|---|---|---|
| 7 | 26014 | Precuneus, posterior cingulate, postcentral gyrus, precentral gyrus, central opercular cortex, lingual gyrus, parahippocampal gyrus, hippocampus, occipital pole, lateral occipital cortex, right Heschl's gyrus, superior temporal gyrus, middle temporal gyrus | -40 | -70 | 34 | 13.9 | < 0.001 |
| 6 | 10980 | Frontal pole, paracingulate gyrus, subcallosal cortex, rostral anterior cingulate, superior frontal gyrus, middle frontal gyrus | -10 | 54 | -4 | 11.8 | < 0.001 |
| 5 | 1247 | Left superior temporal gyrus, middle temporal gyrus, temporal pole | -54 | 0 | -24 | 7.92 | < 0.001 |
| 4 | 598 | Left postcentral gyrus, precentral gyrus, central opercular cortex | -44 | -14 | 36 | 7.35 | 0.001 |
| 3 | 517 | Right superior frontal gyrus, middle frontal gyrus, frontal pole | 24 | 26 | 38 | 7.19 | 0.002 |
| 2 | 333 | Cerebellum | 6 | -50 | -42 | 7.59 | 0.002 |
| 1 | 7 | Cerebellum | 48 | -58 | -40 | 5.52 | 10.951 |

**Table S5. Peak activation for clusters in which HAM-D Improvement was correlated with an increase in negative BOLD signal in the Happy > Fixation contrast. X, Y, and Z coordinates are in MNI space (mm).**

| Cluster Index | Voxels | Brain regions | X | Y | Z | t-value | p-value |
|---|---|---|---|---|---|---|---|
| 5 | 437 | Precuneus | 2 | -74 | 38 | 5.70 | 0.008 |
| 4 | 384 | Left lateral occipital cortex, angular gyrus, supramarginal gyrus | -60 | -58 | 28 | 5.05 | 0.019 |
| 3 | 3 | Left lateral occipital cortex, angular gyrus, supramarginal gyrus | -54 | -58 | 48 | 4.34 | 0.049 |
| 2 | 1 | Cerebellum | -26 | -80 | -28 | 4.81 | 0.049 |
| 1 | 1 | Right lateral occipital cortex | 12 | -80 | 56 | 5.11 | 0.049 |

**Table S6. Peak activation for the clusters from the PPI analysis, in which HAM-D Improvement correlated with an increase in the Happy > Fearful contrast. X, Y, and Z coordinates are in MNI space (mm).**

| Cluster Index | Voxels | Brain regions | X | Y | Z | t-value | p-value |
|---|---|---|---|---|---|---|---|
| 6 | 1115 | Right postcentral gyrus, left and right precentral gyrus, left and right precuneus, left posterior cingulate, right superior parietal lobule, right lateral occipital cortex (superior) | 26 | -24 | 60 | 4.78 | 0.014 |
| 5 | 341 | Left cuneal cortex, lateral occipital cortex (superior), occipital pole | -28 | -74 | 24 | 4.47 | 0.022 |
| 4 | 53 | Right precentral gyrus, middle frontal gyrus | 36 | -2 | 42 | 4.14 | 0.038 |
| 3 | 12 | Left occipital fusiform gyrus | -26 | -68 | -4 | 4.6 | 0.044 |
| 2 | 5 | Left occipital pole | -10 | -94 | 20 | 3.76 | 0.048 |
| 1 | 2 | Left occipital pole | -28 | -90 | 12 | 4.05 | 0.049 |

**Table S7. Regression analysis predicting HAM-D Improvement including change in rostral ACC response (Happy > Fearful) as predictor.**

| | Beta | Standard error | t-value | p-value |
|---|---|---|---|---|
| (Intercept) | -8.92 | 8.4 | -1.06 | 0.2942 |
| Baseline HAM-D | 0.62 | 0.22 | 2.86 | **0.0066** |
| HAM-D change Week 2 | -0.44 | 0.13 | -3.28 | **0.0021** |
| Positive bias change (ACC) | 1.21 | 0.33 | 3.66 | **0.0007** |
| Age | -0.04 | 0.06 | -0.66 | 0.5134 |
| Gender | -0.38 | 1.64 | -0.23 | 0.8187 |
| Medication | -1.37 | 1.23 | -1.11 | 0.2719 |
| TMS Sessions | 0.87 | 0.81 | 1.07 | 0.2894 |

**Table S8. Regression analysis predicting HAM-D Improvement including change in precuneus response (Happy > Fixation) as predictor.**

| | Beta | Standard error | t-value | p-value |
|---|---|---|---|---|
| (Intercept) | -8.78 | 8.20 | -1.07 | 0.2906 |
| Baseline HAM-D | 0.63 | 0.21 | 2.99 | **0.0047** |
| HAM-D change Week 2 | -0.47 | 0.13 | -3.71 | **0.0006** |
| Positive bias change (precuneus) | -1.64 | 0.41 | -4.00 | **0.0003** |
| Age | 0.01 | 0.05 | 0.25 | 0.8032 |
| Gender | -0.10 | 1.59 | -0.07 | 0.9484 |
| Medication | -1.37 | 1.20 | -1.14 | 0.2590 |
| TMS Sessions | 0.57 | 0.79 | 0.72 | 0.4760 |

**Table S9. Regression analysis predicting HAM-D Improvement including change in angular gyrus response (Happy > Fixation) as predictor.**

| | Beta | Standard error | t-value | p-value |
|---|---|---|---|---|
| (Intercept) | -9.93 | 8.12 | -1.22 | 0.2287 |
| Baseline HAM-D | 0.64 | 0.21 | 3.03 | **0.0042** |
| HAM-D change Week 2 | -0.36 | 0.13 | -2.68 | **0.0106** |
| Positive bias change (angular gyrus) | -2.26 | 0.54 | -4.15 | **0.0002** |
| Age | 0.01 | 0.05 | 0.16 | 0.8720 |
| Gender | 2.43 | 1.62 | 1.50 | 0.1409 |
| Medication | 0.16 | 1.17 | 0.14 | 0.8900 |
| TMS Sessions | 0.62 | 0.78 | 0.79 | 0.4317 |

**Table S10. Regression analysis predicting HAM-D Improvement including change in PPI in the precuneus (Happy > Fearful) as predictor.**

| | Beta | Standard error | t-value | p-value |
|---|---|---|---|---|
| (Intercept) | -12.01 | 7.91 | -1.52 | 0.1365 |
| Baseline HAM-D | 0.44 | 0.20 | 2.19 | **0.0345** |
| HAM-D change Week 2 | -0.30 | 0.14 | -2.22 | **0.0322** |
| Positive bias change (PPI precuneus) | 2.89 | 0.63 | 4.57 | **< 0.0001** |
| Age | 0.03 | 0.05 | 0.61 | 0.5477 |
| Gender | 0.45 | 1.51 | 0.29 | 0.7696 |
| Medication | -1.04 | 1.13 | -0.92 | 0.3653 |
| TMS Sessions | 1.39 | 0.77 | 1.80 | 0.0791 |

**Table S11. Regression analysis predicting HAM-D Improvement including change in PPI in the cuneus (Happy > Fearful) as predictor.**

| | Beta | Standard error | t-value | p-value |
|---|---|---|---|---|
| (Intercept) | -5.15 | 8.73 | -0.59 | 0.5585 |
| Baseline HAM-D | 0.48 | 0.22 | 2.16 | **0.0368** |
| HAM-D change Week 2 | -0.43 | 0.14 | -3.11 | **0.0034** |
| Positive bias change (PPI cuneus) | 1.61 | 0.50 | 3.19 | **0.0027** |
| Age | 0.03 | 0.06 | 0.49 | 0.6292 |
| Gender | 0.18 | 1.67 | 0.11 | 0.9133 |
| Medication | -1.08 | 1.25 | -0.86 | 0.3935 |
| TMS Sessions | 0.37 | 0.84 | 0.44 | 0.6604 |

**Table S12. Regression analysis predicting HAM-D Improvement including change in reaction time for happy faces (fMRI gender discrimination task) as predictor.**

| | Beta | Standard error | t-value | p-value |
|---|---|---|---|---|
| (Intercept) | -9.15 | 10.84 | -0.84 | 0.4046 |
| Baseline HAM-D | 0.52 | 0.29 | 1.81 | 0.0790 |
| HAM-D change Week 2 | -0.62 | 0.18 | -3.49 | **0.0014** |
| Positive bias change (RT happy) | 0.00 | 0.01 | 0.44 | 0.6626 |
| Age | 0.01 | 0.07 | 0.12 | 0.9025 |
| Gender | -0.32 | 2.24 | -0.14 | 0.8877 |
| Medication | -0.81 | 1.71 | -0.47 | 0.6409 |
| TMS Sessions | 0.67 | 1.01 | 0.66 | 0.5135 |

**Table S13. Logistic regression analysis predicting Clinical Response including change in Positive Bias Misclassification (FERT) as predictor.**

| | Beta | Standard error | t-value | p-value |
|---|---|---|---|---|
| (Intercept) | -12.03 | 8.00 | -1.50 | 0.1325 |
| Baseline HAM-D | 0.15 | 0.22 | 0.65 | 0.5164 |
| HAM-D change Week 2 | -0.50 | 0.19 | -2.66 | **0.0079** |
| Positive bias change (FERT) | 1.77 | 1.08 | 1.64 | 0.1009 |
| Age | 0.03 | 0.06 | 0.49 | 0.6260 |
| Gender | -0.92 | 1.22 | -0.75 | 0.4523 |
| Medication | 0.25 | 1.03 | 0.25 | 0.8047 |
| TMS Sessions | 0.77 | 0.77 | 1.00 | 0.3180 |
